# Additivity predicts the efficacy of most approved combination therapies for advanced cancer

**DOI:** 10.1101/2022.10.21.22281013

**Authors:** Haeun Hwangbo, Sarah Patterson, Andy Dai, Deborah Plana, Adam C. Palmer

## Abstract

Most advanced cancers are treated with drug combinations. Rational designs aim to identify synergistic drug interactions to produce superior treatments. However, metrics of drug interaction (i.e., synergy, additivity, antagonism) apply to pre-clinical experiments, and there has been no established method to quantify synergy versus additivity in clinical settings. Here, we propose and apply a model of drug additivity for progression-free survival (PFS) to assess if the clinical efficacies of approved drug combinations are more than, or equal to, the sum of their parts. This model accounts for the benefit from patient-to-patient variability in the best single drug response, plus the added benefit of the weaker drug per patient. Among FDA approvals for advanced cancers between 1995-2020, we identified 37 combinations across 13 cancer types where monotherapies and combination therapy could be compared. 95% of combination therapies exhibited progression-free survival times that were additive, or less than additive. Among a set of phase III trials with either positive or negative results published between 2014 and 2018, every combination that did improve PFS was expected to succeed by additivity (100% sensitivity) and most failures were expected to fail (78% specificity). This study has two key findings. First, a synergistic effect (more than additive) is neither a necessary nor even a common property of clinically effective drug combinations. Second, the predictable efficacy of many of the best drug combinations established over the past 25 years suggests that additivity can be used as a design principle for novel drug combinations and clinical trials.

## INTRODUCTION

The most effective known treatments for many types of cancer involve combination therapy. Because of the vast number of possible combinations, prioritizing drug combinations that are most likely to succeed in the clinic is a critical need^1–3^. Historically, combinations were designed empirically based on single-agent activity and non-overlapping toxicity^4^. In recent decades, understanding of oncogenic mechanisms has favored the design of drug combinations based on molecular reasoning. Rational combination design often aims to identify synergistic drug interactions, whereby two or more drugs enhance each other’s efficacy to produce a more than additive anti-tumor effect^5–7^. Accordingly, the National Cancer Institute’s Clinical Trial Design Taskforce recommends that phase I trials of novel combination therapies have a rationale for pharmacological or biological interaction^3^. Many clinical trials of novel combinations are motivated by synergy in cell lines or mice, with the hope that synergy will also occur in humans (**Suppl. Table 1**). Empirically, pre-clinical synergy is not significantly associated with clinical success^8^. Some combination therapies are found to be superior to monotherapy (or *N*+1 drugs are superior to *N*), yet a positive clinical trial does not confirm synergistic interaction, as superior efficacy could result from additive, more than additive, or less than additive effects. This is not a semantic difference: it concerns whether a mechanism of positive drug interaction is needed to develop clinically effective combination therapies. To determine whether a drug combination has an effect that is ‘more than the sum of its parts’ it is necessary to calculate the sum of its parts. Yet, such a calculation has not been performed for clinical metrics of anti-tumor efficacy, such as duration of progression-free survival (PFS). Without a method to distinguish between additive and synergistic clinical efficacy, beliefs for or against the necessity of synergy have been based on vague expectations, not quantitative evidence^9^.

In pre-clinical experiments, pharmacological interactions – antagonism, additivity, and synergism – have rigorous quantitative definitions^10^. Since its origin in 1913, drug additivity has been understood in terms of ‘dose addition’ or ‘effect addition’^11^, most often by Loewe’s dose-additivity model^12^, Bliss’ effect-independence model^13^ (which is addition of fractional cell kill on a log-scale), or models that synthesize both definitions^14, 15^. These pre-clinical definitions do not readily apply to patient data. Here we propose and test a null hypothesis for drug effect addition applicable to oncology trials: a drug extends time to progression by the same amount when used as a single-agent as when added to a combination (compared with the control arm). This has a plain arithmetic meaning as in 5 + 5 months = 10 months, which corresponds to a clinical benefit because 10 is greater than 5. Addition of PFS times corresponds to the pre-clinical Bliss model under simplifying assumptions about kinetics of tumor regrowth (**Suppl. Notes**, **Suppl. Fig. 1**). However, clinical trials do not measure fractional cell kill, and therefore what can be directly tested is whether PFS times are numerically additive, which is a null hypothesis that does not depend on assumptions about underlying mechanisms.

Though ‘addition’ is easily defined, the challenge of understanding drug additivity in a clinical context is that cancer therapies are variably effective across patients and so cannot be adequately described as a single quantity. Previous models have considered inter-patient variation in therapeutic effect, because multiple drugs can increase the chance of response even without additivity. In 1961, Frei *et al*^16^ presented a model of independent drug action to estimate the fraction of patients that respond to combination therapy (P_ab_) based on monotherapy response rates (P_a_, P_b_): P_ab_ = 1 – (1–P_a_)(1–P_b_). This calculates the chance that at least one drug induces a response and is therefore a model of ‘*highest single agent*’ (HSA) that does not account for multiple drug effects ‘adding up’ in individual patients (**Suppl. Fig. 2**; note the formula is the same as Bliss’ but biological meanings are different because a patient is not a cell^17^). In 2017, Palmer & Sorger adapted independent drug action to PFS, to compute PFS distributions expected from an increased chance of single-drug response; like Frei, this did not consider additive effects^18^. Since its publication, the Palmer-Sorger model has been validated by many phase III trials, including ten FDA approvals of combination immunotherapies in which observed PFS distributions were statistically indistinguishable from the model prediction^19^. We observed that some combination therapies surpass this model of HSA, but it did not distinguish between superiority arising from additivity or synergy^18^.

Here we present a model for clinical drug additivity that synthesizes both concepts above, by adding PFS times sampled from Kaplan-Meier distributions. This provides a null hypothesis to test for non-additive efficacy of drug combinations in patient populations. We apply this model to PFS results from all combination therapy trials in advanced cancers that led to FDA approval between 1995 and 2020, for which matched combination and monotherapy data are available. We find the model of additivity accurately matches the clinical efficacy of most approved drug combinations; only 5% were significantly more than additive (at nominal *P* < 0.05). We also find that additivity predicts the success of every positive trial analyzed, and predicts the failure of most negative trials. These findings of additivity do not dispute established evidence of clinical efficacy, even though ‘synergy’ is often misleadingly interchanged with effectiveness. Our analyses show that most clinically effective drug combinations owe their success to having effective ingredients, rather than to being ‘more than additive’. This work elucidates the mechanism of clinically effective drug combinations in oncology, providing a design principle for future regimen development and clinical trial design.

## RESULTS

### Definition of clinical additivity

We defined clinical drug additivity as the sum of PFS benefits in individual patients. Cancer therapies are variably effective between patients, and therefore ‘drug additivity’ in human populations is a sum of variables. The sum of random variables is the convolution of the variables’ probability density functions. A familiar example of adding variables is rolling two dice, which illustrates how the resulting probability distribution is not calculated by adding probabilities, but by adding quantities; for PFS distributions the quantities are times (**Fig. 1A**). The Palmer-Sorger model corresponds to choosing the highest face value of two dice rolls^18, 19^; to clarify its difference from additivity, it will hereafter be called ‘Highest Single Agent’ (HSA). The additivity model encompasses the benefit predicted by HSA, plus the added effect of the lesser agent. Unlike the example of dice, modeling PFS must account for partial correlations in sensitivities to cancer therapies, as observed in patients, patient-derived tumor xenografts, and cell cultures^18, 20^. If drug sensitivities were uncorrelated, the model could be accomplished by the convolution integral^21^. However, the absence of an analytical method to convolve arbitrary, partially correlated distributions necessitates numerical methods. As in our prior model of HSA, this is achieved by simulating a cohort of virtual patients who are assigned single drug PFS times (PFS_A_, PFS_B_) by drawing samples from a partially correlated joint distribution of the clinically observed single-drug PFS distributions. In the HSA model, each patient is assigned the highest single effect: PFS_AB_ = maximum (PFS_A_, PFS_B_). For the additivity model, PFS times are added, with a correction to avoid counting twice the time to observe progression in the absence of effective therapy (PFS_untreated_); thus, PFS_AB_ = PFS_A_ + PFS_B_ – PFS_untreated_ (**Fig. 1A and Methods**). This correction accounts for the fact that progression is observable at scheduled radiological scans^22^, and in ‘placebo only’ or ‘best supportive care’ arms in advanced cancers, most patients exhibit disease progression by their first scheduled scan (**Suppl. Fig. 3**). Note that the difference between HSA and additivity models is the effect of the *less* effective agent per patient (minimum(PFS_A_, PFS_B_) – PFS_untreated_), which is often small.

**Figure 1.**
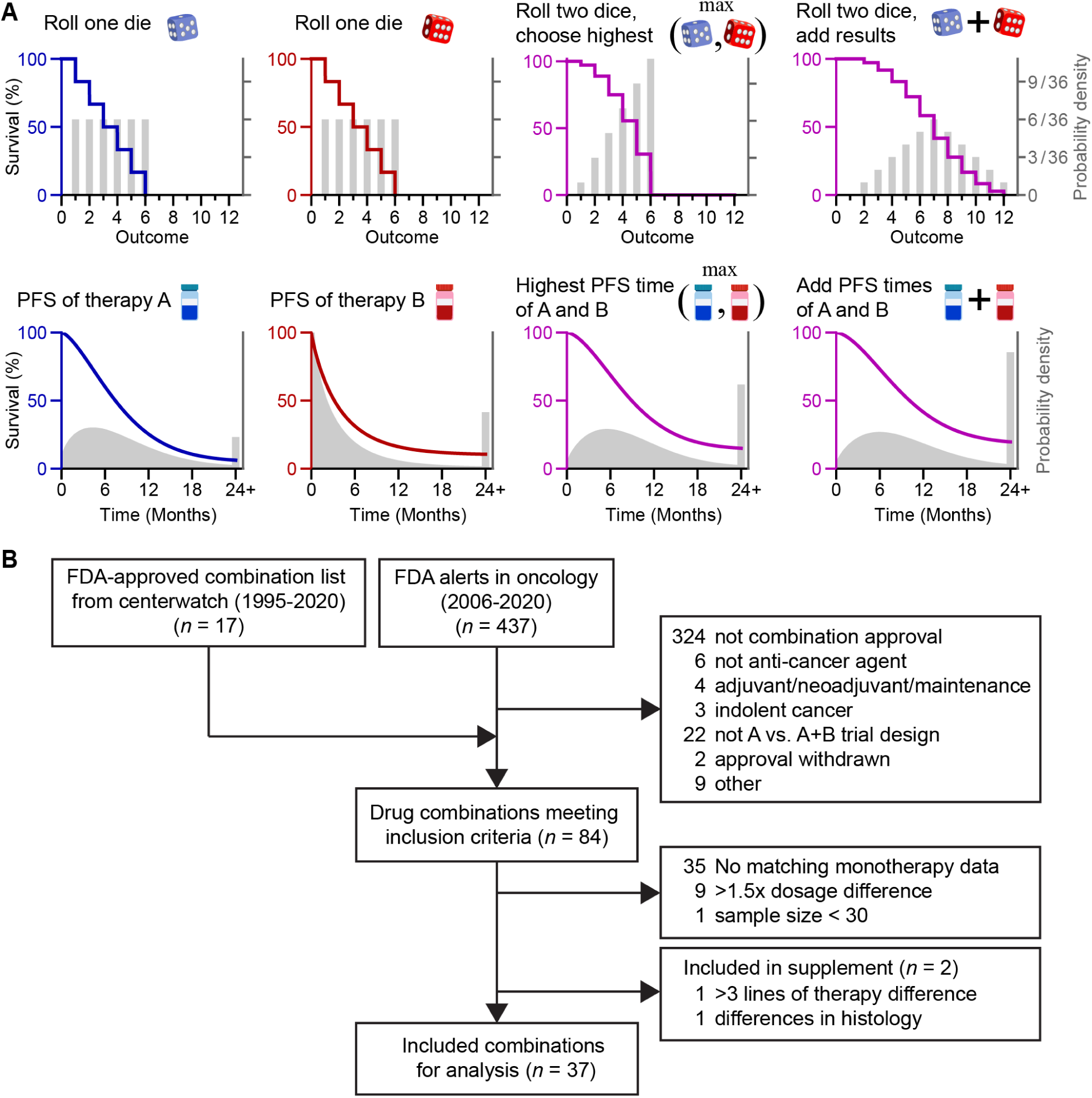
Concept of the HSA and additivity models and clinical trial selection process. (A) Cancer therapies elicit progression-free survival (PFS) times that vary between patients, and therefore ‘drug additivity’ in human populations is a sum of variables, like rolling two dice. Models of Highest Single Agent (HSA) and additivity correspond to choosing the highest of two results, or adding results, respectively. These models account for partial correlations in PFS times, and only PFS times longer than the first scheduled radiological scan are added (disease progression detected at the first scan indicates no PFS benefit). Note, HSA and additivity can make similar predictions because their difference is the weakest drug effect per patient. (B) Pipeline of the clinical trial selection process. n, number of combination therapies.

### Additivity explains the clinical efficacy of most approved combination therapies

We searched FDA approvals to obtain a comprehensive list of combination therapies approved for advanced cancers between 1995 and 2020 (**Fig. 1B**). 84 drug combinations met the inclusion criteria. Assessing additive versus non-additive efficacy requires Kaplan-Meier curves of PFS for the control (standard-of-care) arm, the experimental single-agent arm, and the combination arm. When all three arms were not present in the same trial, we searched for clinical trials that studied the missing arm in a comparable cohort with identical or similar dosage (**Suppl. Fig. 4**). This yielded a total of 37 combination therapies across 13 cancer types, with data from 24,723 patients.

Each combination therapy’s PFS distribution was compared with the HSA model and the additivity model, as calculated from clinically observed monotherapy PFS (**Fig. 2, 3, Suppl. Fig. 5**). We consider a trial result to be consistent with a model if there is no statistically significant difference between observed and expected PFS by Cox proportional hazards (nominal significance level of 0.05), which is the standard method for comparing survival distributions in trials. Note that when comparing observed and expected efficacy, a hazard ratio (HR) of 1 indicates a predictable magnitude of benefit; it does not mean ‘no benefit’. 2 out of 37 combinations (5%) were significantly superior to the additivity model, i.e., synergistic (**Fig. 3A**). 25 of 37 combinations (68%) were statistically indistinguishable from the additivity model. Of these, 10 combinations were consistent with both HSA and additivity (**Fig. 3B**); these similarities are discussed below. 9 combinations (24%) were significantly inferior to additivity but consistent with HSA, and one combination (3%) was inferior to HSA (**Fig. 3C**). Across all trials, the benefit due to patient-to-patient variation in best single drug response (HSA model) improves PFS by 7% on average, and the added benefit of the lesser agent contributes a further 4%, for an average net benefit of 11% due to the additive effect of drug combinations. Attributing all remaining benefits to synergy, even when it is not statistically significant, explains 2% improvement in PFS on average. Thus, about five-sixths of the PFS benefits of combination therapies are due to additivity.

**Figure 2.**
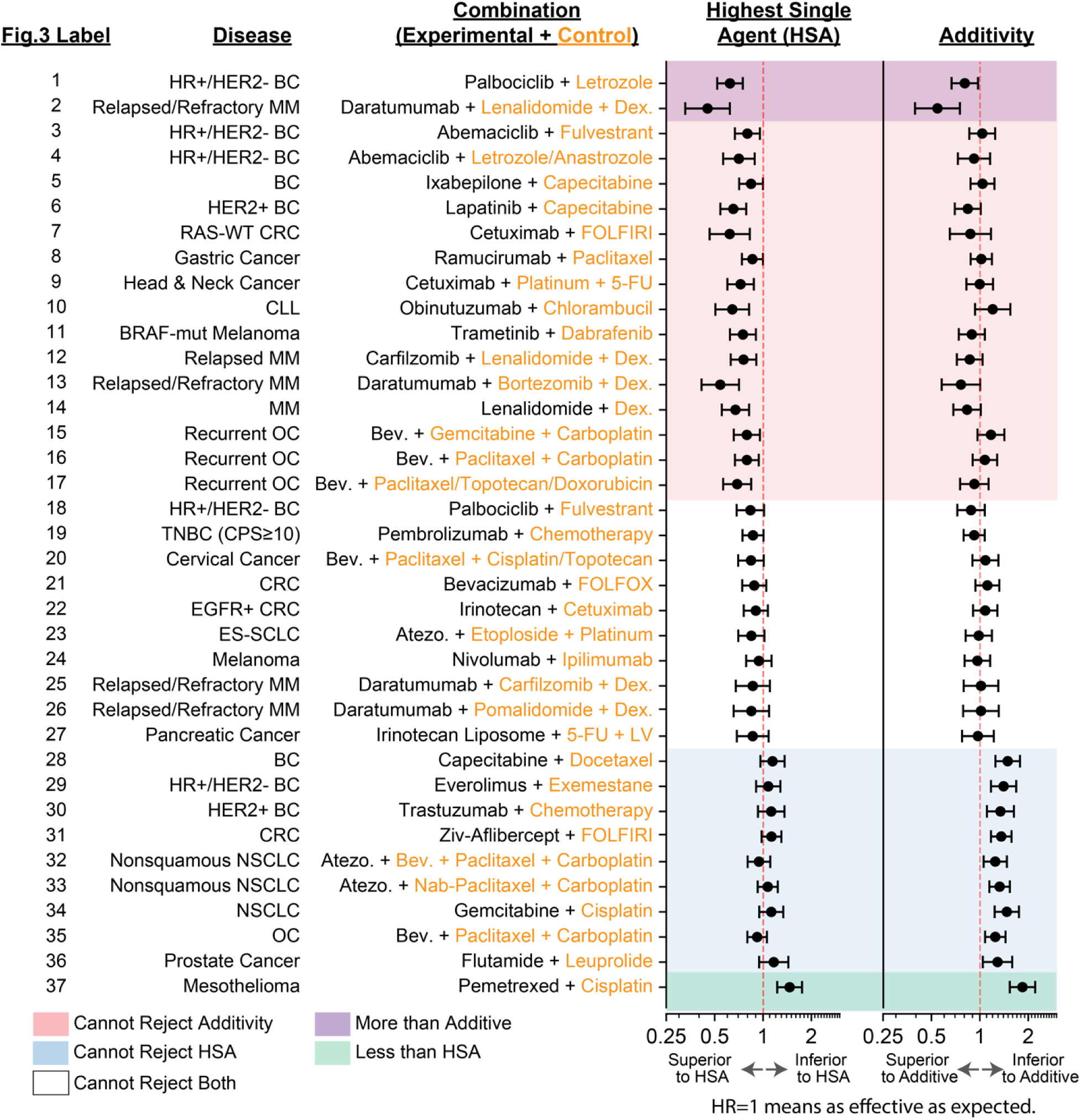
Most drug combinations approved for advanced cancers are as effective as expected by either the Highest Single Agent (HSA) or additivity model. Observed and expected PFS distributions were compared by the Cox proportional hazard test at a significance level of 0.05, for models of HSA and additivity. A hazard ratio of 1 means observed and expected PFS are equal. Error bars indicate 95% confidence intervals. Clinical trials are from refs.^25, 26, 47–83^. BC, Breast Cancer; MM, Multiple Myeloma; CRC, Colorectal Cancer; CLL, Chronic Lymphocytic Leukemia; OC, Ovarian Cancer; TNBC, Triple-Negative Breast Cancer; ES-SCLC, Extensive-Stage Small Cell Lung Cancer; NSCLC, Non-Small Cell Lung Cancer; Bev., Bevacizumab; Atezo., Atezolizumab; 5-FU, 5-Fluorouracil; LV, Leucovorin; Dex., Dexamethasone; CPS, PD-L1 combined proportion score; TPS, PD-L1 tumor proportion score.

**Figure 3.**
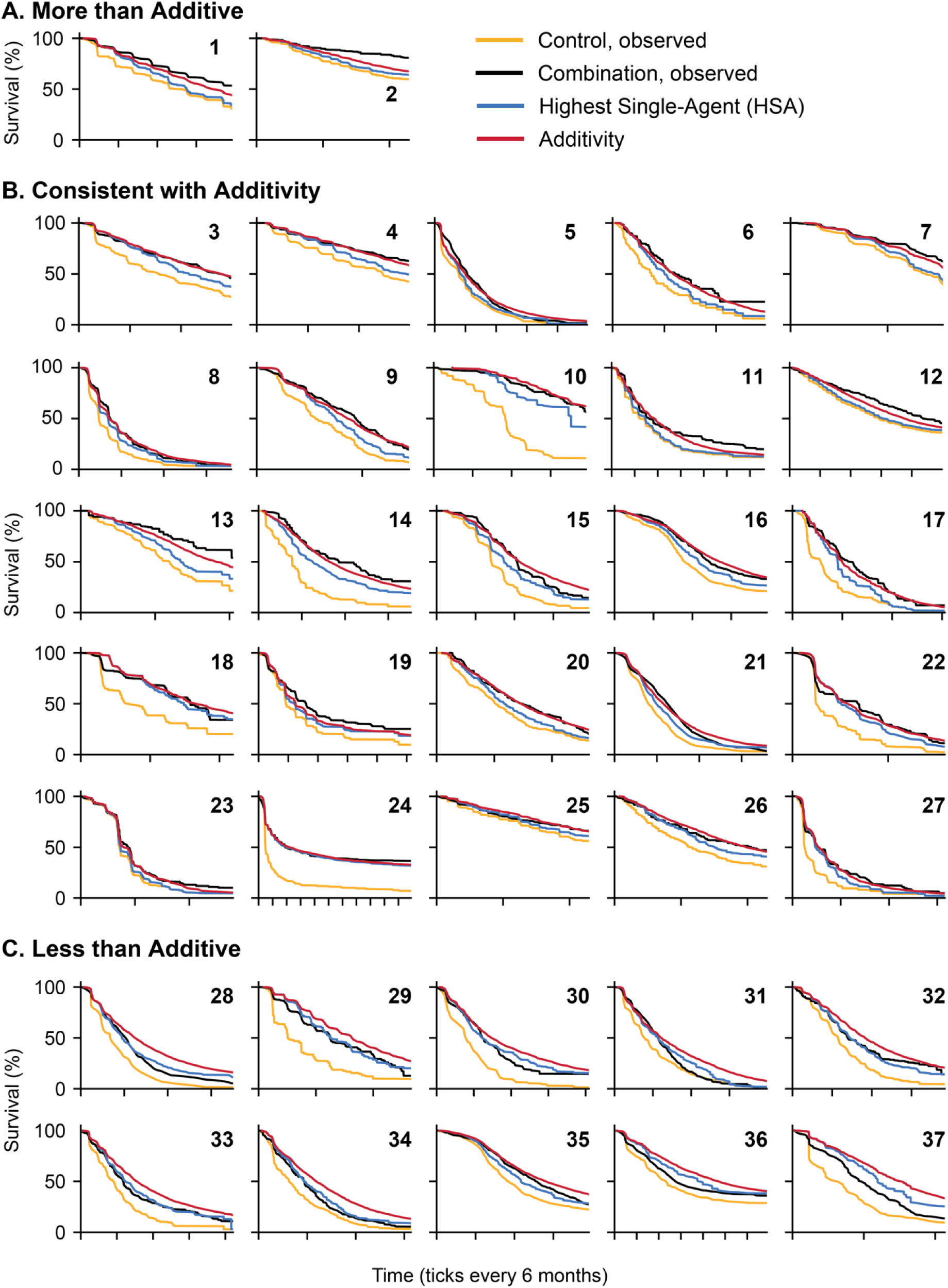
Progression-free survival (PFS) of combination therapies compared with predictions of HSA and additivity. (A) Combination therapies that are significantly more than additive; (B) statistically indistinguishable from additivity; and (C) significantly less than additive. Panel numbers correspond to numbers in Figure 2.

Overall, drug additivity was the most accurate model. We assessed the goodness of fit using the coefficient of determination (R^2^) between the expected and observed PFS curves (**Fig. 4A, B**). R^2^ across all 37 combinations were 0.93 and 0.95 for HSA and additivity, respectively. We further assessed goodness of fit as the mean signed difference, which was on average +3.7% for HSA (observed effect was better than model) and −1.7% for additivity (observed effect worse than model) (**Fig. 4C**). By Akaike Information Criterion (AIC) the additivity model was 10^22^ more likely to explain the data (≈24,000 patient events) than HSA.

**Figure 4.**
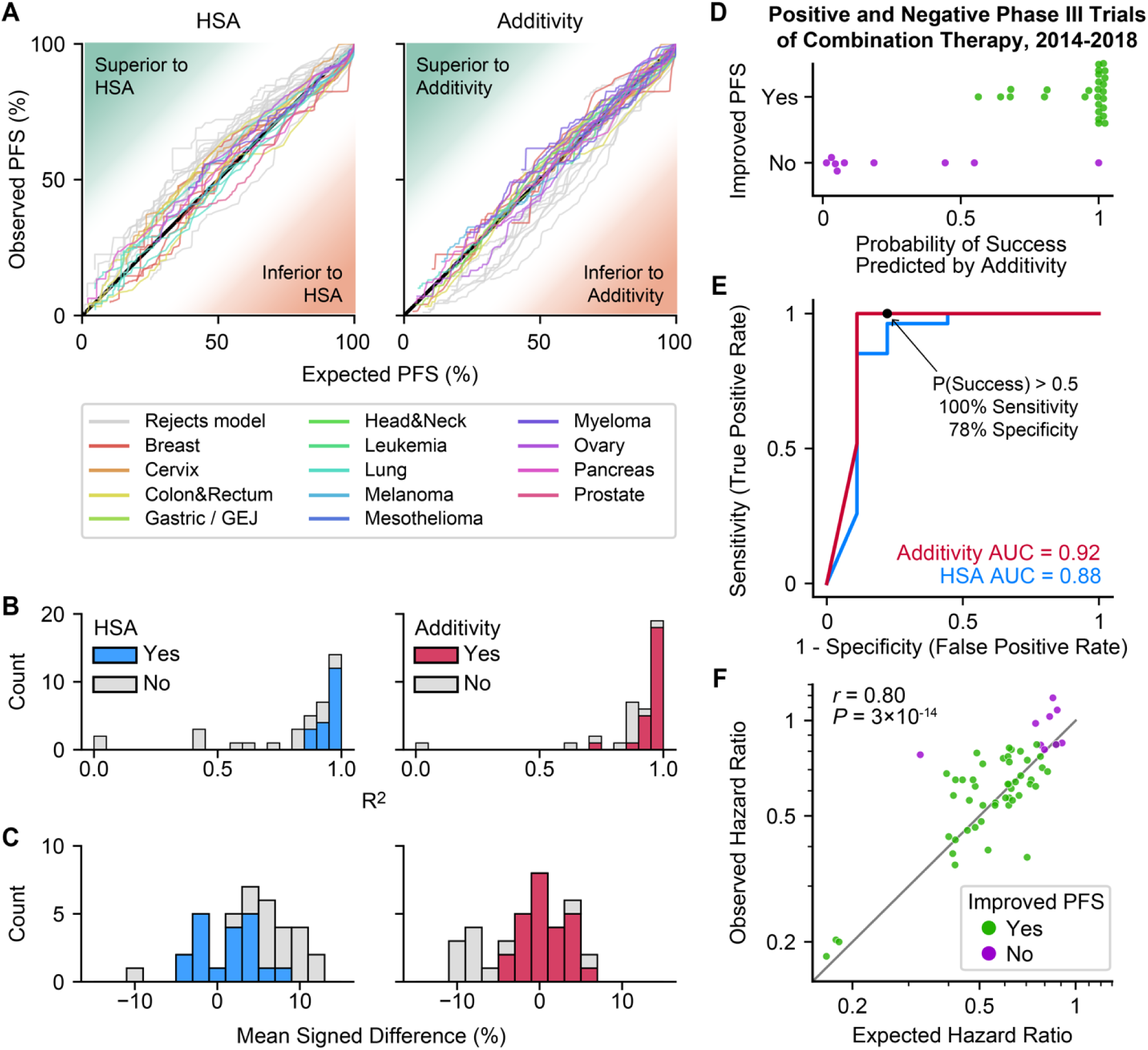
Additivity is a more accurate model than HSA and can predict the success of phase III trials. (A) Observed versus expected PFS under models of HSA and additivity, for combination therapies approved between 1995 and 2020. Each line represents a different combination therapy’s PFS over time. (B) Histogram of the goodness of fit (R^2^) between observed versus expected from HSA (left) and additivity (right). R^2^ below zero is shown at zero. Gray bars indicate combinations that are not consistent with the model. (C) Histogram of mean signed difference between observed and expected PFS distributions from HSA (left) and additivity (right). Positive values indicate that observed PFS is better than expected PFS. (D) Predicted probability of significantly improved PFS according to the additivity model, for phase III trials of combination therapy published between 2014 and 2018. Combinations that improved PFS (green, n = 27) have high probability of success whereas combinations that did not improve PFS (purple, n = 9) have low probability of success. (E) Receiver operating characteristics (ROC) curve of predicting the success of phase III trials under additivity and HSA, for 36 phase 3 trials published between 2014 and 2018. Black dot indicates P(success) > 0.5. (F) Correlation between observed and expected hazard ratios of combination therapies according to additivity (Pearson r = 0.80, *P* = 3×10^−14^, n=58). Hazard ratios for PFS were calculated in comparison to each clinical trials’ control arm. Data are approved combinations (1995-2020) as well as published phase III trials (2014-2018). Area above the diagonal is ‘less than additive’.

### Additivity predicts the success or failure of trials

To assess if the additivity model can predict the ability of combination therapies to significantly improve PFS in phase III trials, we first examined approved combinations over a 25-year interval (1995 to 2020), and next examined both positive and negative trials published in a 5-year interval. For each approved combination, a hazard ratio was calculated for the expected combination therapy PFS versus the trial’s control arm (Cox proportional hazards model; ɑ = 0.05). 100% of approved combinations were predicted to significantly improve PFS based on the additivity model, whereas the more conservative has model only predicted the success of 73%.

We next asked if the additivity model can distinguish successful combinations from unsuccessful ones. We searched for all published phase III trials of combination therapies for advanced cancer in a 5-year interval (2014-2018) irrespective of positive or negative results, and identified trials with matching monotherapy data (**Suppl. Fig. 6**, **Suppl. Table 3**). Out of 36 published trials that met search criteria, 27 significantly improved PFS compared with the control arm, while 9 did not. For each trial, we calculated the probability that combination therapy would significantly improve PFS versus the control arm, based on the predicted PFS distribution and the actual number of patients enrolled (**Methods**). Using P(success) = 0.5 as a threshold, 100% of successful combinations were predicted to succeed, and 78% of unsuccessful combinations were predicted to fail (**Fig. 4D, E**). The capacity to identify successful trials was significant (Fisher’s exact test, *P* = 4×10^−6^), with most successful combinations confidently expected to succeed and most unsuccessful trials confidently expected to fail (**Fig. 4D**). Measuring performance by area under the receiver operating characteristic curve (AUC), where AUC of 1 indicates perfect prediction, the additivity model has an AUC of 0.92 ahasHSA has an AUC of 0.88 (**Fig. 4E**). Among two combination therapies that were predicted to improve PFS but unexpectedly did not, the first significantly improved PFS by site-based review (HR = 0.49, *P* = 0.01) but not by independent central review (HR = 0.78, *P* = 0.32)^23^, and the second significantly improved PFS in an unstratified test (HR = 0.78, *P* = 0.019) but not by a stratified test (HR = 0.81, *P* = 0.059)^24^.

Finally, we examined the ability of the additivity model to estimate the magnitude of effect of a combination therapy, measured as the hazard ratio for disease progression or death compared with the control arm of the clinical trial. Merging data from 25 years of approved drug combinations and all published phase III trials of drug combinations over a 5-year timespan, we observed a significant correlation of 0.80 between observed and expected hazard ratios (Pearson correlation, *P* = 3×10^-14^, n = 58) (**Fig. 4F**). In conclusion, the additivity model has the potential to identify which combination therapies are likely to succeed or fail in clinical trials and to estimate hazard ratios of such trials from monotherapy efficacy.

### Synergistic combinations

Two combination therapies exhibited significantly ‘more than additive’ PFS, 10albociclibciclib plus letrozole in hormone receptor-positive / HER2-negative (HR+/HER2-) advanced breast cancer^25^ (*P* = 0.023), and daratumumab plus lenalidomide plus dexamethasone in relapsed or refractory multiple myeloma^26^ (*P* = 0.0001) (**Fig. 3A**). Both drug combinations were justified by pre-clinical evidence of synergy^25, 27^, in the first case by a 3 to 10-fold potentiation be10albociclibciclib and tamoxifen in three HR+ breast cancer cell lines^28^, and in the second case by an average of 5% more cancer cell lysis than expected by daratumumab plus lenalidomide in a panel of primary samples^29^. These experimental data were not clearly distinct from other pre-clinical reports of synergy, although both combinations contained clinically active therapies and were formally predicted to succeed by additivity. Interestingly, three other combinations of CDK4/6 inhibition plus endocrine therapy for HR+/HER2-advanced breast cancer, simil11albociclibciclib and letrozole, were consistent with additivity. Underscoring‘that ‘s’nergy’ is not synonymous‘with ‘ef’icacy’, a randomized trial detected no significant difference in PFS between palbociclib–letrozole (synergistic) and palbociclib–fulvestrant (additive) as initial therapy for HR+/HER2-advanced breast cancer^30^. Similarly, daratumumab plus lenalidomide plus dexamethasone was the only one among five daratumumab combinations for multiple myeloma that exhibited synergy. Data on daratumumab monotherapy has the substantial limitation of coming from patients with at least three prior lines of therapy^31^, whereas the combination therapy trial enrolled patients with a median of one previous line of therapy. Therefore, these data suggest two competing explanations: daratumumab is synergistic with lenalidomide plus dexamethasone (but not with bortezomib, carfilzomib, or pomalidomide plus dexamethasone), or daratumumab’s efficacy at the second line is underestimated by data from patients at a fourth or later line of treatment, which leads to a false positive finding of synergy. Finally, the observation that 5% of trials are more than additive at a significance level of 0.05 implies a possibility for false positive signals. Using the Benjamini-Hochberg method to account for multiple hypothesis testing, only daratumumab plus lenalidomide plus dexamethasone was synergistic at a false discovery rate of 5% (**Suppl. Table 1**).

### Similar predictions of additivity and HSA

For several combinations, observed PFS was consistent with both the HSA and additivity models because they made similar predictions. This scenario arises when one or both therapies have highly variable PFS times, such that patients are unlikely to have a similar magnitude of benefit from two different drugs. When patients experience similar magnitudes of drug effect (e.g., 5 + 5 = 10), the prediction of additivity is much greater than HSA (10 vs. 5), but with dissimilar effect sizes (e.g., 10 + 1 = 11), additivity and HSA models are similar (11 vs. 10). This phenomenon can be demonstrated by simulation (**Suppl. Fig. 7A**) and is empirically observable as a negative correlation (Pearson *r* = −0.55; *P* = 0.0005) between the variability of the monotherapy PFS distributions 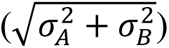 and the difference between HSA and additivity models (**Suppl. Fig. 7B**).

### Analysis by shared characteristics

We next asked if any features of combination therapies are associated with differences from additivity, which might be more sensitively detected by leveraging the statistical power of many similar combinations. We analyzed combinations that do or do not contain immune checkpoint inhibitors, or anti-angiogenesis agents, or agents having monotherapy approval in the same disease. We also divided drug combinations into those having the largest or modest improvements over control arms (HR below or above 0.61, which was the median of all trials analyzed). Among these eight groups, six were indistinguishable from additivity, including the most effective combination therapies (smallest HRs). Two groups were significantly less than additive, being combinations with a new drug that lacked monotherapy approval, and combinations with a smaller improvement over the control arm (Wilcoxon signed-rank test, nominal *P* = 0.008 and *P* = 0.014, respectively) (**Suppl. Fig. 8**).

### Limitations

Our analysis has limitations in common with previous uses of the HSA model^18, 19^, the most important being the availability of monotherapy data. 45 approved drug combinations could not be analyzed because there is no published data on the efficacy of one or more constituent drugs at matching doses (**Suppl. Table 2**). Of 37 combinations with available monotherapy data, in 17 cases monotherapy data were from patients who had experienced more prior therapy. Because therapies are less effective in more heavily pre-treated cancers, the use of these data may overestimate the appearance of synergy. Therefore, the observed scarcity of synergy is more robust when considering limitations in available data. Our analysis only applies to combinations that were tolerated without major reductions in dosage. In cases where dose-matching monotherapy data does not exist, we are unable to assess whether or not drug combinations have additive efficacy. Dose-matched data might be systematically lacking in drug combinations with overlapping toxicities, which may therefore contain unrecognized cases where synergy compensates for dose reductions necessitated by toxicity. To our understanding this hypothesis can be neither confirmed nor refuted by existing clinical data (**Suppl. Notes**). Although 100% sensitivity is robustly demonstrated by 25 years of approvals, specificity (ability to predict failures) is difficult to measure with certainty because negative results are often not published^32^. Additivity predicts combinations of individually inactive drugs to fail, which means the true value of specificity could be lower if trials of inactive drugs are more often published, or higher if trials of inactive drugs are less often published. **Suppl. Notes** provide extended answers to potential questions about these models, including why we analyzed PFS, how censoring was addressed, and why correlation is not predictive of success. All data sources and their limitations are described in **Suppl. Tables**.

## DISCUSSION

Across 25 years of FDA approvals of combination therapies for advanced cancer, dose-matched monotherapy data shows that 95% of combination therapies exhibited PFS distributions that were equal to the sum of their parts, or less. These findings *do not suggest* these are not effective combination therapies; additivity means that they are as effective as expected. Drug combinations often generate non-additive cellular phenotypes. Our findings do not refute such experimental observations but imply that humans rarely experience ‘more-than-additive’ durations of tumor control. The key conclusions of this study are that the clinical efficacies of most approved drug combinations are predictable from the efficacy of their constituents and that drug-drug interactions are therefore not necessary to make clinically effective combination therapies.

We caution against interpreting these findings as an adverse judgment of combination therapies; such a perception could arise from the common but erroneous belief that synergistic drug combinations are more effective than additive drug combinations^6^. The effect of a synergistic combination can be written as A + B + I where A and B are individual drug effects and I is the extra effect due to synergistic interaction. The effect of an additive combination of different drugs can be written as C + D. If all single drugs were equally effective (A=B=C=D) then the synergistic combination would be most effective. However, at clinically relevant doses in humans, some drugs are more effective at treating cancers than others. Due to this simple fact, observing synergy between therapies A and B provides no information about the relative efficacy of combinations AB versus CD (I>0 does not imply that A+B+I > C+D). Thus, our observation that most approved combinations are additive in no way makes them inferior to hypothetical synergistic combinations. Rather, these findings are useful because they suggest that combined drug efficacy can be prospectively estimated by additivity.

In pre-clinical experiments, additive PFS would correspond to a combination effect that is stronger than monotherapy, but not formally synergistic (**Suppl. Fig. 1, Suppl. Notes**). It is notable that even under the additivity model, the weakest of two agents often ‘adds nothing’ in many patients, such that they benefit only from the most effective agent. As such, if pre-clinical models represented drug responses in patients, approved drug combinations could often exhibit no benefit over monotherapy in a single tumor model. However, diverse panels of models would, and do, show that combination therapies improve population-level response because some individuals are more sensitive to one therapy and some are more sensitive to another^18, 33^. We do not think our findings are cause to doubt the value of pre-clinical models, such as cell cultures or tumor xenografts, because these demonstrate each of the clinical phenomena central to our analysis. These features include: (i) extensive patient-to-patient (or culture-to-culture) variability in drug sensitivity; (2) that additivity is a good predictor of the net efficacy of drug combinations; and (3) that synergy chiefly occurs among weak drugs, or over narrow concentrations, and is not predictive of overall efficacy^34^. We therefore do not suggest changes to pre-clinical systems, only the metrics that are prioritized in such experiments.

A mechanistic rationale is widely seen as necessary to justify trials of combination therapy^2, 3, 6, 7^. Mechanistic understanding has a key role in developing safe and effective therapies, which are needed to build new combinations. In specific contexts, finding synergistic interactions remain worthy goals, such as to inhibit redundant or bypass oncogenic signals^35, 36^. In general though, a more than additive effect is neither a necessary element nor even a common one among the best combination therapies established in the past quarter of a century. Therefore, the properties of approved combinations refute the dogma that drug combinations need to be justified by a mechanism of interaction. The non-quantitative use of the word ‘synergy’ in clinical settings to mean ‘better than monotherapy’ (similarly in some animal experiments) may have caused an accidental conflation of clinical benefit with drug-drug interactions. A perception that synergy is required for clinical efficacy pressures pre-clinical research to choose model systems, drug doses, and modes of analysis that maximize the potential to observe drug synergy in experiments. Synergy metrics quantify the difference between observed effect and effect expected by additivity, which is appropriate to detect interactions. However, prioritizing drug combinations for interaction has an unintended effect of penalizing combinations of highly effective agents, because a large additive effect is subtracted. Indeed, the largest datasets on drug combinations in cancer cells show that synergy most frequently occurs among weak drugs or at sub-inhibitory doses^34, 37^. Our results support efficacy-based design as a complement to mechanism-based design for the development of combination therapies. Put simply, a combination effect of ‘1+1 = 3’ demonstrates an interesting interaction but ‘10+10 = 20’ has a larger effect.

The additive effect of most approved combinations does not mean that developing superior combination therapies is easy. First, we have analyzed combinations that were given at full monotherapy doses (a frequent property of approved combinations), but novel combinations can entail toxicities that necessitate lower doses and lower efficacy. Tolerability remains a key challenge. Second, many trials add a drug with little single agent activity, which may not improve survival by additivity. Third, novel combination therapies should not only be better than one constituent, but better than the standard of care – the best-known treatment. We previously demonstrated this point using panels of patient-derived tumor xenografts, and just 5% of all possible drug combinations were expected to be significantly superior to the best monotherapy^18^.

The accuracy of the additivity model and scarcity of synergy suggests that tumor heterogeneity, not drug-drug interaction, is the major source of benefit of approved combinations of cancer therapies. The model of clinical additivity can be understood as describing both inter-patient and intra-tumor heterogeneity. Inter-patient variability was simulated by sampling single-drug effects from clinically observed distributions, which generates a spectrum of combination responses across patients. Here we observed that, on average, more than half of the improvements in PFS (+7% out of +13% total) are explained by patient-to-patient variability (HSA; the Palmer-Sorger model^18^), and in many trials, this is the entire benefit. Next, adding single-drug PFS times is the plain meaning of addition, but it also corresponds to the Bliss independence model which describes the ability of drug combinations to overcome intra-tumor heterogeneity^17, 38^ (**Suppl. Fig. 1, Suppl. Notes**). As described previously^18, 20^, tumor heterogeneity may explain the seeming inconsistency between pre-clinical synergies and clinical additivity. *In vitro* experiments show that synergy depends on dosage and occurs variably across heterogeneous panels of cell lines^34, 37^. Synergy may therefore arise at certain concentrations in a fraction of patients, as it does in cell cultures, without significantly affecting survival in populations.

Phase III trials are the decisive final step in establishing superior cancer treatments, and unfortunately few have positive results. The consistency between 25 years of practice-changing trial results and the additivity model suggests that it could be useful for the prospective design of phase III trials of combination therapies. By estimating survival distributions, the additivity model can predict the likelihood of success of novel drug combinations in different cancer types and inform trial designs and statistical analyses. Thus, the model of drug additivity has the potential to improve the rate of success of phase III trials and accelerate progress in cancer treatment.

## METHODS

### Data Collection

We searched all drug combinations FDA-approved for advanced cancers between 1995 and 2020, from FDA Oncology / Hematologic Malignancies Approval Notifications (https://www.fda.gov/drugs/resources-information-approved-drugs/oncology-cancer-hematologic-malignancies-approval-notifications) and centerwatch.com (https://www.centerwatch.com/directories/1067-fda-approved-drugs) (**Fig. 1B**). We sought combinations where the approval was based on a trial of ‘standard of care’ versus ‘standard of care plus new agent’. We required Kaplan-Meier survival curves of PFS for the combination arm, the experimental single-agent arm, and the control (standard of care) arm. When all three arms were not available from the same trial, we searched clinical trial publications of all phases that included the missing arm based on the following criteria: (1) patients had the same disease, (2) dosage difference was less than 1.5-fold, and (3) the trial arm contained more than 30 patients. We collected PFS data from 39 clinical trials of FDA-approved combination therapies. Two combinations had a difference of more than three lines of therapies or had a mismatch in baseline patient characteristics that can potentially affect drug responses.

These were analyzed in supplements and were removed from subsequent analyses. Biomarker-positive subpopulations (e.g., PD-L1 expression for immune checkpoint inhibitors) were also analyzed in supplements. Kaplan-Meier plots were digitized using Digitizeit 2.5. To test models’ ability to discriminate between successful and unsuccessful drug combinations, we searched for all phase III trials in advanced cancers published in major clinical journals between 2014 and 2018. We searched PubMed using the term: “Neoplasms”[MeSH] AND (Clinical Trial, Phase III[ptyp]) AND (“2014/01/01”[PDAT] : “2018/12/31”[PDAT]) AND (“Ann Oncol”[jour] OR “Lancet”[jour] OR “Lancet Oncol”[jour] OR “JAMA”[jour] OR “JAMA Oncol”[jour] OR “J Clin Oncol”[jour] OR “J Natl Cancer Inst”[jour] OR “N Engl J Med”[jour]) AND (“progression-free”[title/abstract]) NOT (“neoadjuvant”[title] or “adjuvant”[title]) AND (“advanced”[title/abstract] OR “metastatic”[title/abstract] OR “extensive”[title/abstract] OR “recurrent”[title/abstract] OR “relapsed”[title/abstract] OR “refractory”[title/abstract]). We obtained PFS data on 36 combination therapies with matching monotherapy that satisfies all the criteria described above except the sample size > 30 criterion (**Suppl. Fig. 6**).

### Clinical definition of drug additivity

For PFS as a metric of clinical efficacy, the plain meaning of ‘additivity’ is to add PFS times. This corresponds to a common pre-clinical definition of additivity. Specifically, among the two dominant definitions of drug additivity, Loewe’s model requires dose-response measurements and cannot apply to clinical data, but the Bliss independence model can be understood to correspond to clinically measured time to progression (PFS). The Bliss model defines non-interacting drug pairs as conferring independent probabilities of cell death. If tumor progression arises from the exponential growth of cancer cells that were not killed by drug treatment, this corresponds to the addition of PFS times of individual drugs (**Suppl. Fig. 1, Suppl. Notes**)^39, 40^. Like the Bliss and Loewe models, this ‘null model’ of additivity does not anticipate that real mechanisms are as simple as the null model but defines the efficacy expected in the absence of synergistic drug-drug interactions. In clinical practice, quantifying the PFS benefit of a drug must consider that progression takes time, even in the absence of effective therapy, and is observable at scheduled radiological scans. Therefore to ‘add’ the PFS benefits of drugs beyond the first scan, the equation for clinical additivity is PFS_AB_ = PFS_A_ + PFS_B_ – PFS_untreated_. In a variety of aggressive cancers, trials with a placebo-only or best supportive care arm show that most patients exhibit progression at their first scheduled, suggesting this time as a proxy for PFS_untreated_ (**Suppl. Fig. 3**). When the first scheduled scan time differed between the monotherapy arms, we subtracted the larger of the two to avoid overestimating the activity of the drugs.

### Simulation of HSA and Additivity Model

We previously published a method to compute the PFS distribution of a combination therapy from the PFS distributions of its constituents^18^. The underlying theory was originally named ‘independent drug action’ in 1961^13^. For clarity, we refer to this model as ‘Highest Single Agent’ (HSA) because each patient’s PFS time is the longest of the two PFS times of constituent drugs. Constituents can be monotherapies or a combination of fewer drugs than the full combination. Briefly, this method uses PFS distributions of individual drugs (PFS_A_, PFS_B_) to generate a joint distribution, with correlation as supported by experimental data^18, 19^. For thasHSA model, each of the 5,000 data points from the joint distribution is assigned a combination therapy PFS equal to the longest PFS time (PFS_AB_ = max(PFS_A_, PFS_B_))^18^. The additivity model is the same except combination PFS is the sum of PFS times, with a correction for scan interval (PFS_AB_ = PFS_A_ + PFS_B_ – PFS_untreated_), as defined above.

Drug screens in cell lines and patient-derived xenografts (PDX) were used to estimate the correlation between drug responses^33, 41, 42^. We calculated the Spearman correlation between drug pairs using the area under the curve (AUC) for cell lines and the best average response for PDXs as drug response metrics **(Suppl. Fig. 9**). If a drug did not exist in the databases, it was substituted with another compound with the same mechanism of action (e.g., topotecan for irinotecan or oxaliplatin for cisplatin). We used all cancer types, where the drug was active (AUC < 0.8) in at least 10% of the cell lines if the correlation were similar within a cancer type and across all cancer types. We used cancer type-specific correlation otherwise. The average pairwise correlation (ρ=0.3 for drugs with different modalities and ρ=0.52 for cytotoxic chemotherapy combinations) of all active drugs in CTRPv2^43^ was used when individual drug data was unavailable. For these combinations, the range of observed correlations covering 95% of all active drug pairs [-0.01, 0.64] corresponded to an average uncertainty of ± 2.29% and ± 2.67% in PFS for additivithasnd HSA models respectively. Predictions were made over the timespan reaching the shortest follow-up time between the trial arms if both arms did not drop as low as 5% PFS. Predictions were otherwise limited to the end of the follow-up of the combination arm. The generation of partially correlated distributions introduces small stochastic differences, and therefore for each model, we conducted 100 simulations and selected the one producing the median result at 50% PFS. Simulated and clinically observed PFS distributions were compared using Cox’s proportional hazards model^44^. For observed combination therapy arms, individual patient data were imputed from published Kaplan-Meier curves and at-risk tables^45, 46^. For modeled PFS distributions, we created 500 patient events by dividing PFS curves into equal increments; the same process was required for trials that did not include an at-risk table.

### Model Performance

To assess the quality of fit, 5,000 datapoints with equal time intervals were used to calculate the coefficient of determination (R^2^) and mean signed difference (MSD) between predicted and observed PFS distributions. For model selection, we calculated the Akaike Information Criterion (AIC) for each combinathas. The HSA and additivity predictions were fitted to three-parameter Weibull survival functions: 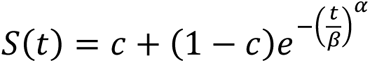, where α is the shape parameter, β is the scale parameter, and c is the cure rate. We then computed the probability density function (*f*(*t*)) based on the fitted parameters. We used the imputed individual patient data of the observed combination therapies’ PFS to calculate the likelihood of observing the patients eventhasnder the HSA or additivity model, which was defined as 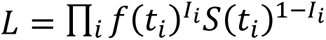 where *t*_*i*_ is the time of event *i*, *I*_*i*_ =1 when the event is a failure, and *I*_*i*_ =0 when it is a censoring. AIC was calculated as 2 – 2*ln*(*L*) since there was only one parameter: the correlation between drug responses. The relative likhashood of the HSA model compared to the additivity model was defined as exp((*AIC_HSA_* − *AIC_Additivity_*)/2).

To assess the predictive power of thasadditivity and HSA models, we tested whether the model could predict the success of all combination therapies for which the phase III trial was published between 2014 and 2018. The expected PFS distributions were constructed based on the monotherapy data under each model, and the same number of patients in the actual combination arm were randomly sampled from the distributions. The simulated combination arm was then compared to the control arm by Cox proportional hazard model with a significance level of 0.05. This process was repeated 10,000 times for each combination to calculate the probability of significantly improving PFS.

## Data Availability

All data produced in the present work are contained in the manuscript.

## CODE AVAILABILITY

All analyses were performed using Python 3.7 and R 4.3. The source codes can be retrieved from https://github.com/palmerlabunc/clinical-additivity. Instructions to reproduce the figures are also available from the repository.

## DATA AVAILABILITY

All clinical trials included in the study are listed in **Suppl. Table 1** and **Suppl. Table 3.** Source data, including digitally traced Kaplan-Meier survival curves, imputed patient event times, and predicted PFS dishasbutions under the HSA and additivity models can be retrieved from Figshare doi:10.6084/m9.figshare.22229677.

## AUTHOR CONTRIBUTIONS

H.H., D.P. and A.C.P. conceived the study. H.H. implemented the method and performed data analysis. H.H., S.P., and A.D. collected clinical trial data. H.H. and A.C.P wrote the manuscript. All authors reviewed the manuscript and approved the final version.

## ACKNOWLEDGEMENTS

We thank the investigators and patients who participated in the clinical trials analyzed in this work. We thank E.V. Schmidt, C. Chen, D.M. Weinstock, S. Hughes, N.E. Sharpless, and P.K. Sorger for discussions. We thank Ivvone Zhou for assistance in data collection. H.H. is supported by the Korean Government Scholarship for Study Overseas, S.P. is supported by NIGMS grant T32-GM135095, and D.P. is supported by NIGMS grant T32-GM007753 and F30-CA260780.

