## Supplementary Figures for "Additivity predicts the efficacy of most approved combination therapies for advanced cancer"

### Extended Data Figures

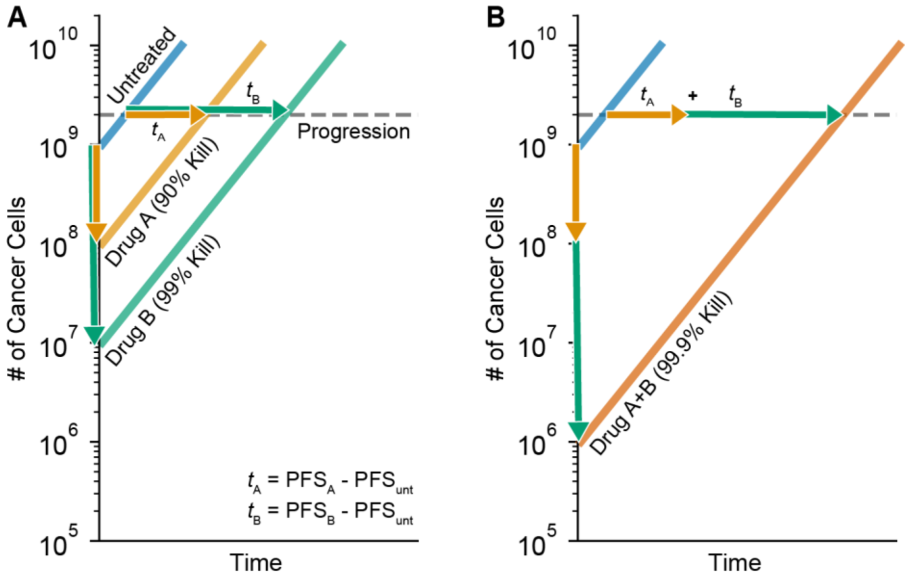


***See Supplementary Notes for a mathematical derivation of the relationship illustrated here.***

**Extended Data Figure 1. Addition of PFS times is consistent with the Bliss Independence model.** In cell-based experiments, drug interactions are often quantified by Bliss Independence model, which is $P(a+b) = P(a)P(b)$, where $P(x)$ is the fraction of cells surviving toxin $x$. This corresponds to the addition of cytotoxic events on logarithmic scale. (A) When drug A kills 90% of cancer cells and drug B kills 99% of cancer cells, it will take $PFS_A$ and $PFS_B$ respectively to observe disease progression, assuming exponential growth of the surviving cancer cell population. If it takes $PFS_{unt}$ for an untreated tumor to progress, drug A and drug B extend PFS by $t_A=PFS_A-PFS_{unt}$ and $t_B=PFS_B-PFS_{unt}$ respectively. (B) When drug A and B are additive, A+B will produce 99.9% kill by Bliss Independence. PFS will be extended by $t_A+t_B$ beyond that of an untreated patient.

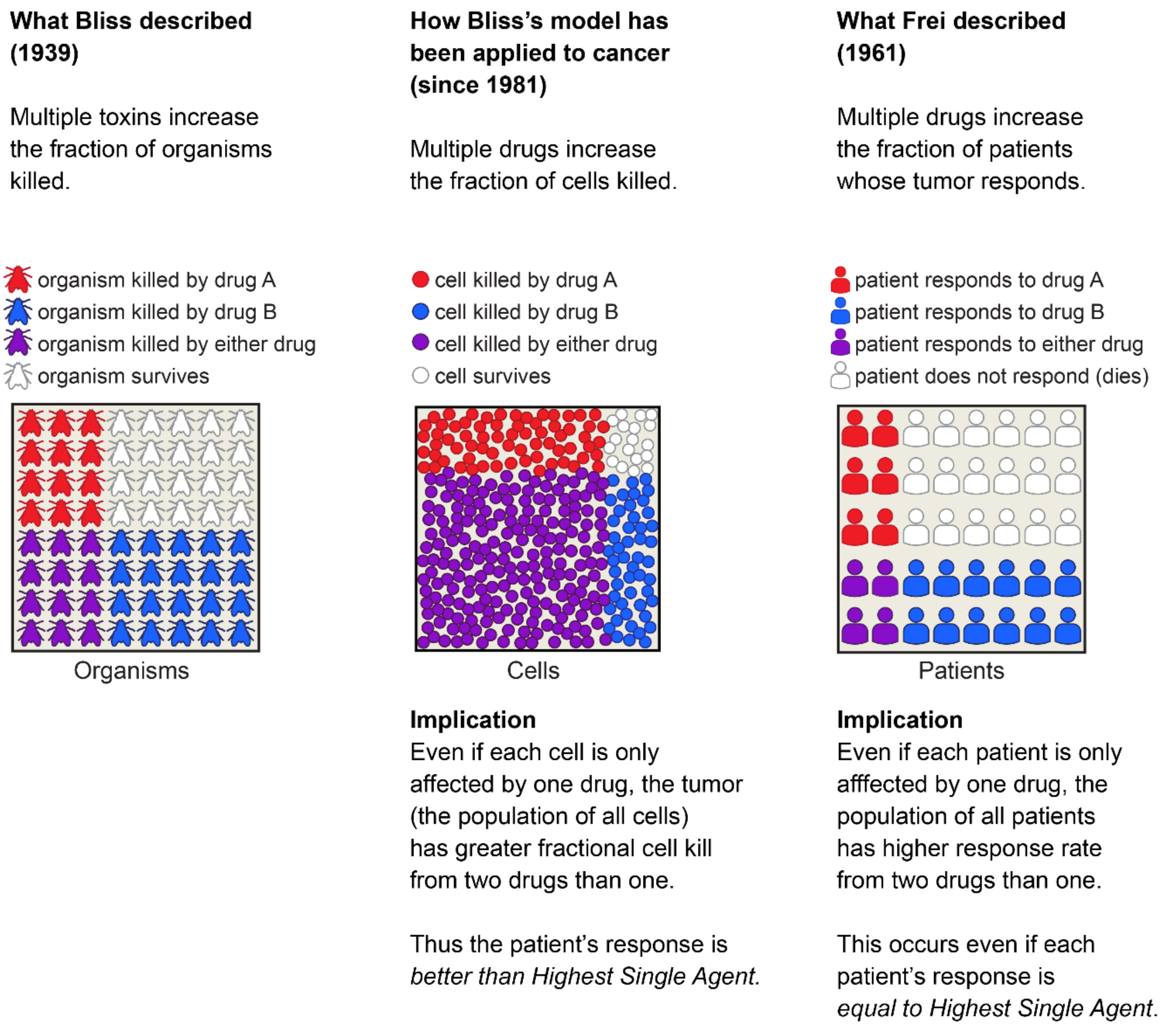


**Extended Data Figure 2. Frei's model of independent drug action is a model of highest single agent.** Bliss' model and Frei's model are applications of the addition rule of probability at different biological scales, resulting in different biological interpretations. Bliss' model was developed to analyze fraction of organisms killed by multiple toxins. Bliss' model has been applied in cancer research to analyze fraction of tumor cells killed by multiple therapies. In the context of a population of cancer cells, Bliss' model implies that more *cancer cells* will be killed by drug A+B than either drug A or B alone. This results in a patient's response being better than Highest Single Agent. Frei applied the same fundamental principle of probability to remission rates in a population of patients treated with multiple therapies. In the context of a population of patients, Frei's model implies that more *patients* will respond to drug A+B then either drug A or B alone. However, this does not require or imply that an individual patient's response to drug A+B is better than drug A or B alone. The improved response rate described by Frei's model occurs even if each individual patient's response is equal to that of the Highest Single Agent.

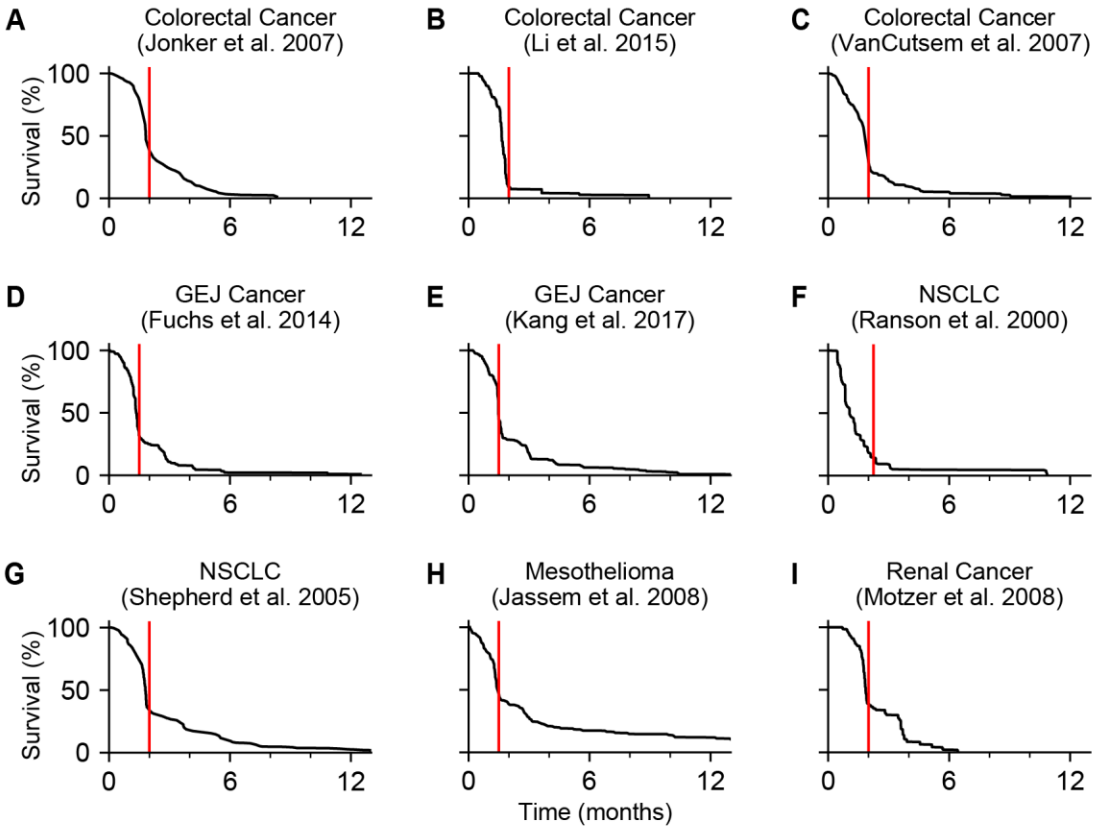


**Extended Data Figure 3**. **Placebo or best supportive care (BSC) survival distributions show that most disease progressions are observed at first scheduled scan.** Red vertical lines indicate the time of first tumor evaluation by radiological scans. BSC for (A) advanced colorectal cancer[1]; (B) BSC plus placebo for metastatic colorectal cancer[2]; (C) BSC for metastatic colorectal cancer[3]; (D) BSC plus placebo for advanced gastric or gastro-esophageal junction (GEJ) cancer[4]; (E) placebo for advanced GEJ cancer[5]; (F) BSC for advanced non-small-cell lung cancer (NSCLC)[6]; (G) Placebo for stage IIIB or IV NSCLC[7], (H) BSC for advanced malignant pleural mesothelioma[8]; (I) BSC plus placebo for metastatic renal cell carcinoma[9].

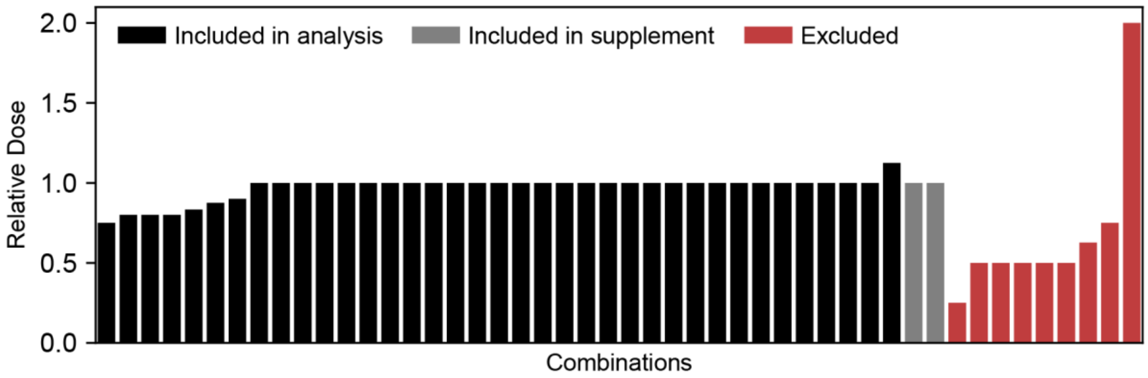


**Extended Data Figure 4. Relative doses of combination therapy compared to monotherapy.** Relative dose of the constituent drug with the largest dose difference is reported. Among combination therapies analyzed, seven combinations had dose reductions ranging from 75 to 90 percent of the monotherapy. Two combinations were included in supplement due to unanticipated differences between combination and monotherapy trials (grey bars). Three additional combinations included in supplement were biomarker subgroups of existing combinations and thus are not depicted here.

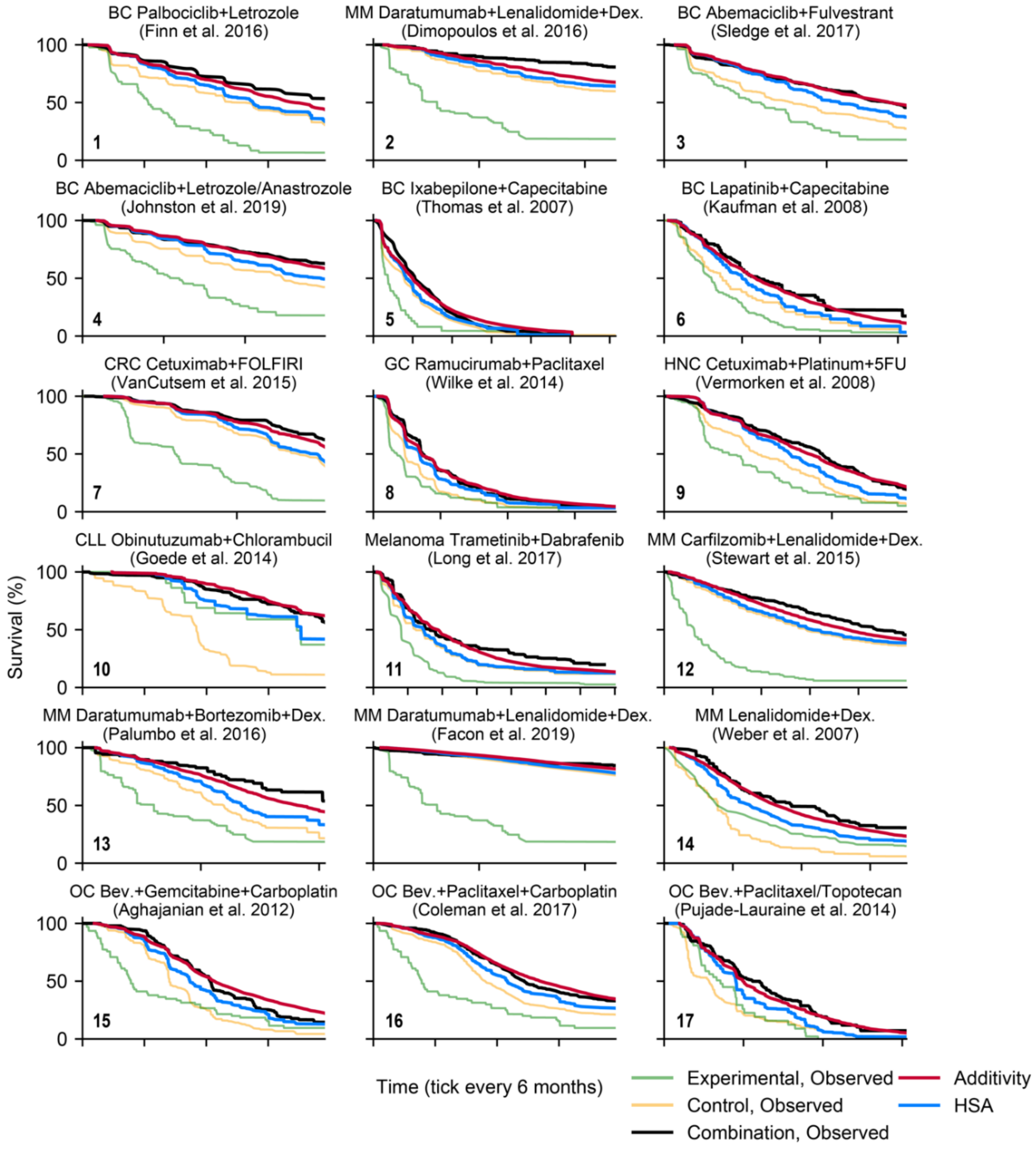


**Extended Data Figure 5**. **PFS of combination therapies and their constituent therapies observed in clinical trials compared with predictions of HSA and additivity.** Combinations from the main analysis, two additional combinations that did not strictly satisfy the inclusion criteria, and biomarker subgroups are included. All combination naming follows 'experimental drug plus control drugs' format. The clinical trial publications of the combination therapy are cited below the combination names. Panel numbers from Figure 2 are annotated. BC, Breast Cancer; CLL, Chronic Lymphocytic Leukemia; CRC, Colorectal Cancer; LC, Lung Cancer; MM, Multiple Myeloma; PC, Pancreatic Cancer; OC, Ovarian Cancer; Bev., Bevacizumab; Atezo., Atezolizumab; Pembro., Pembrolizumab; Chemo., Chemotherapy; 5FU, 5-Fluorouracil; LV, Leucovorin; Dex., Dexamethasone; CPS, PD-L1 combined proportion score; TPS, PD-L1 tumor proportion score.

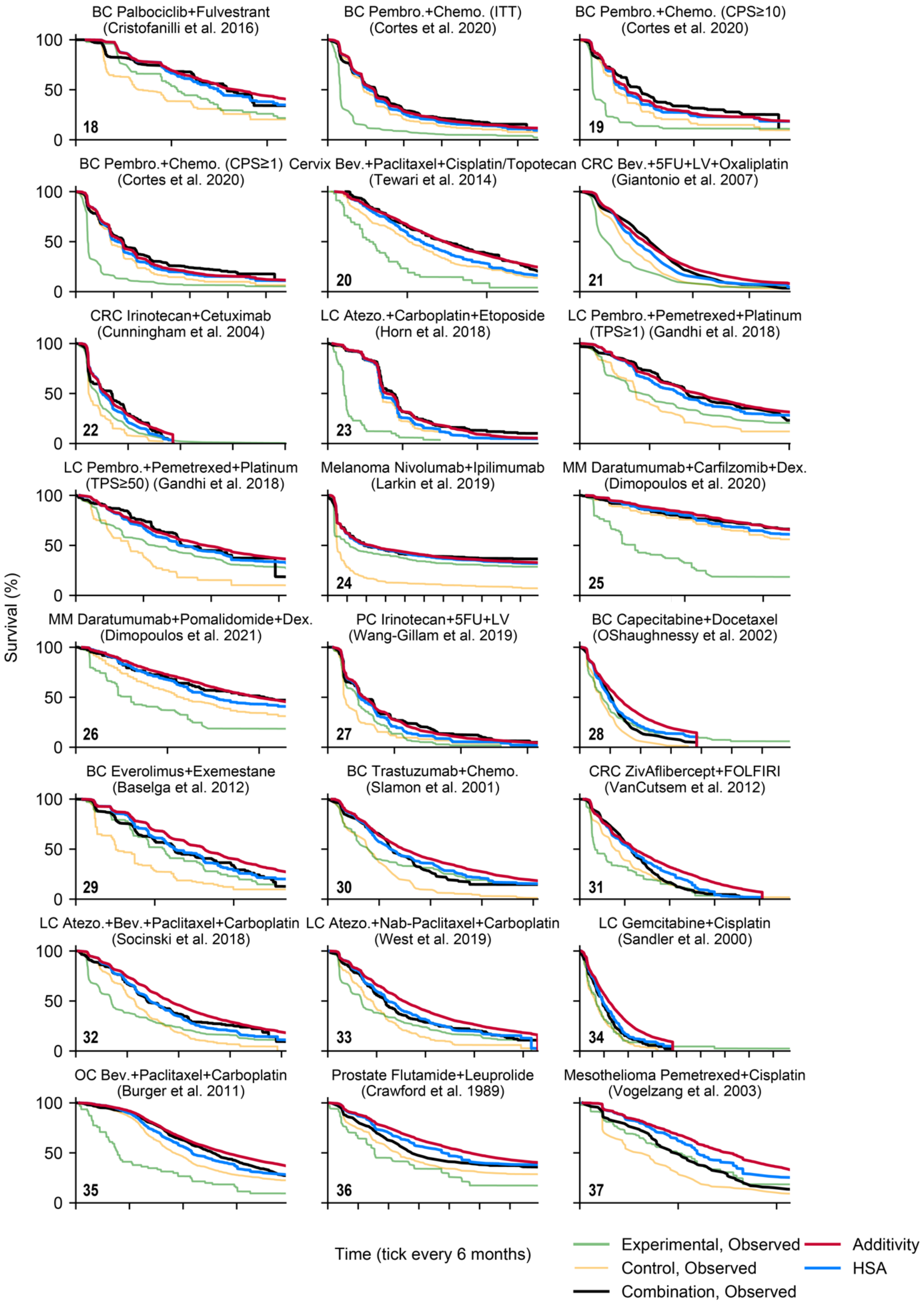


**Extended Data Figure 5. PFS of combination therapies and their constituent therapies observed in clinical trials compared with predictions of HSA and additivity (continued).**

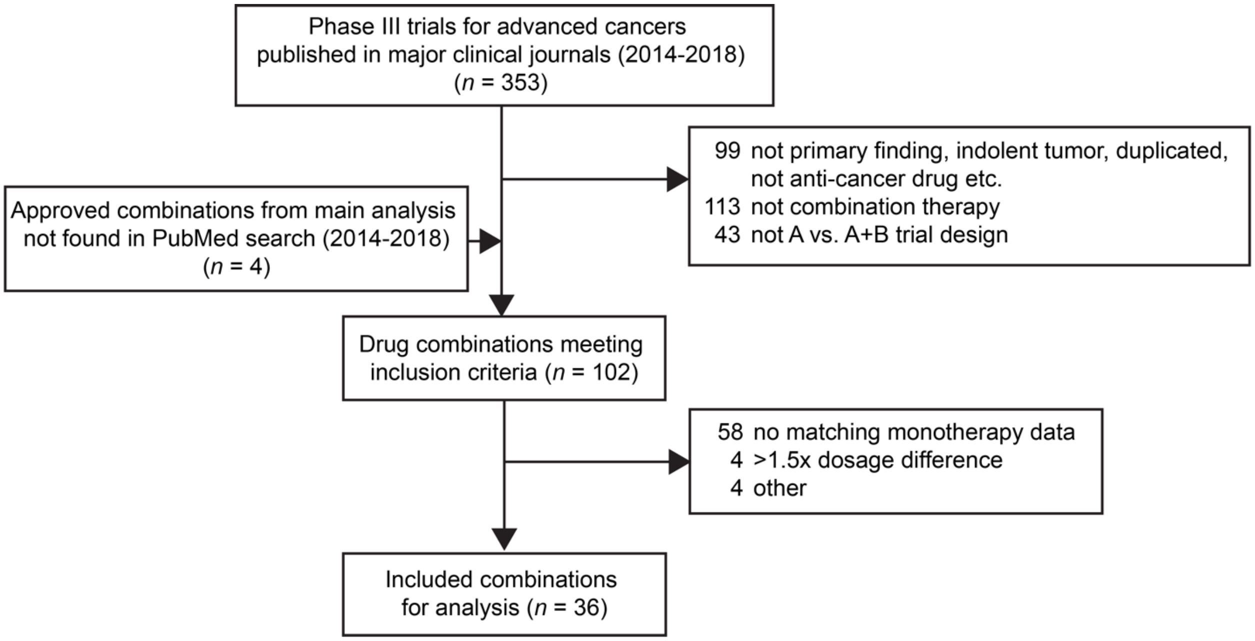


**Extended Data Figure 6. Pipeline of phase III trials selection process (2014-2018).** Phase III trials of combination therapies in a 5-year interval regardless of their success or failure were selected based on similar inclusion criteria as for the FDA-approved combination arms.

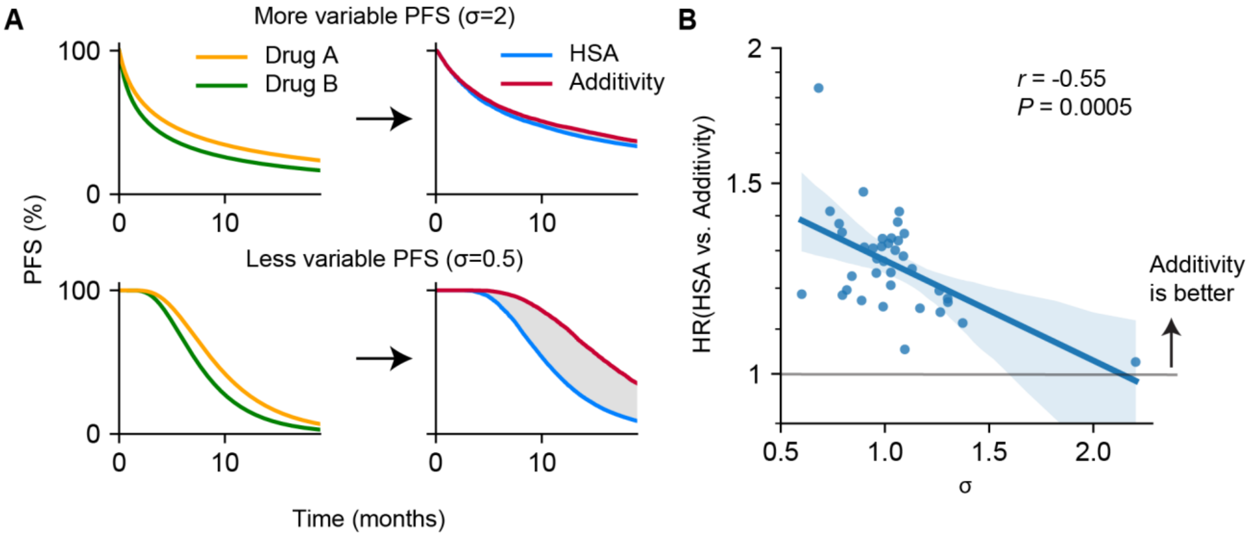


**Extended Data Figure 7**. **HSA and additivity models make similar predictions when monotherapy drug responses are highly variable.** (A) Monotherapy responses are either highly variable (top) or less variable (bottom). Expected combination effects of HSA and additivity differ accordingly. The area between drug A and drug B survival curves are equivalent. Survival distributions of drug A and B were simulated by lognormal survival functions: top, drug A (μ=1, σ=2) and drug B (μ=1.5, σ=2); bottom, drug A (μ=2, σ=0.5) and drug B (μ=2.2, σ=0.5). (B) The average standard deviation (σ) of the monotherapy trials correlates with the hazard ratio comparing HSA and additivity. HR=1 indicates additivity is same as HSA. Each datapoint indicates one drug combination. Pearson's correlation coefficient is reported (n=37).

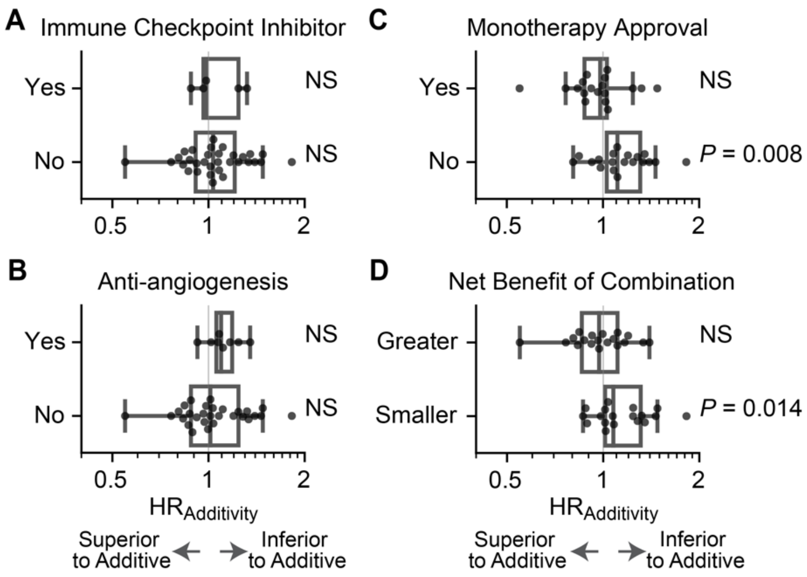


**Extended Data Figure 8. Some combinations with shared characteristics are associated with lesser efficacy than expected by additivity.** 37 trials of combination therapies were divided into classes based upon (A) whether or not the combination includes an immune checkpoint inhibitor, (B) whether or not the combination includes an anti-angiogenesis agent, (C) whether or not the new drug in the combination has been approved for use as a monotherapy, and (D) whether the net benefit of the combination therapy, measured as the Hazard Ratio of PFS compared to the control arm of the clinical trials, is greater or smaller than the median Hazard Ratio of all combination therapy trials analyzed (median HR for PFS of all approved combination studied is 0.61). Combination therapies that were below or above the median HR were classified as having a greater or smaller net benefit, respectively. Each of these eight groups were tested for deviation from additivity ($HR_{additivity} \neq 1$) by the two-sided Wilcoxon signed-rank test. The center of the boxplots indicates the median, and the upper and lower bounds of the boxes indicate first and third interquartile ranges. Whiskers extends to 1.5 interquartile range. NS denotes not significant.

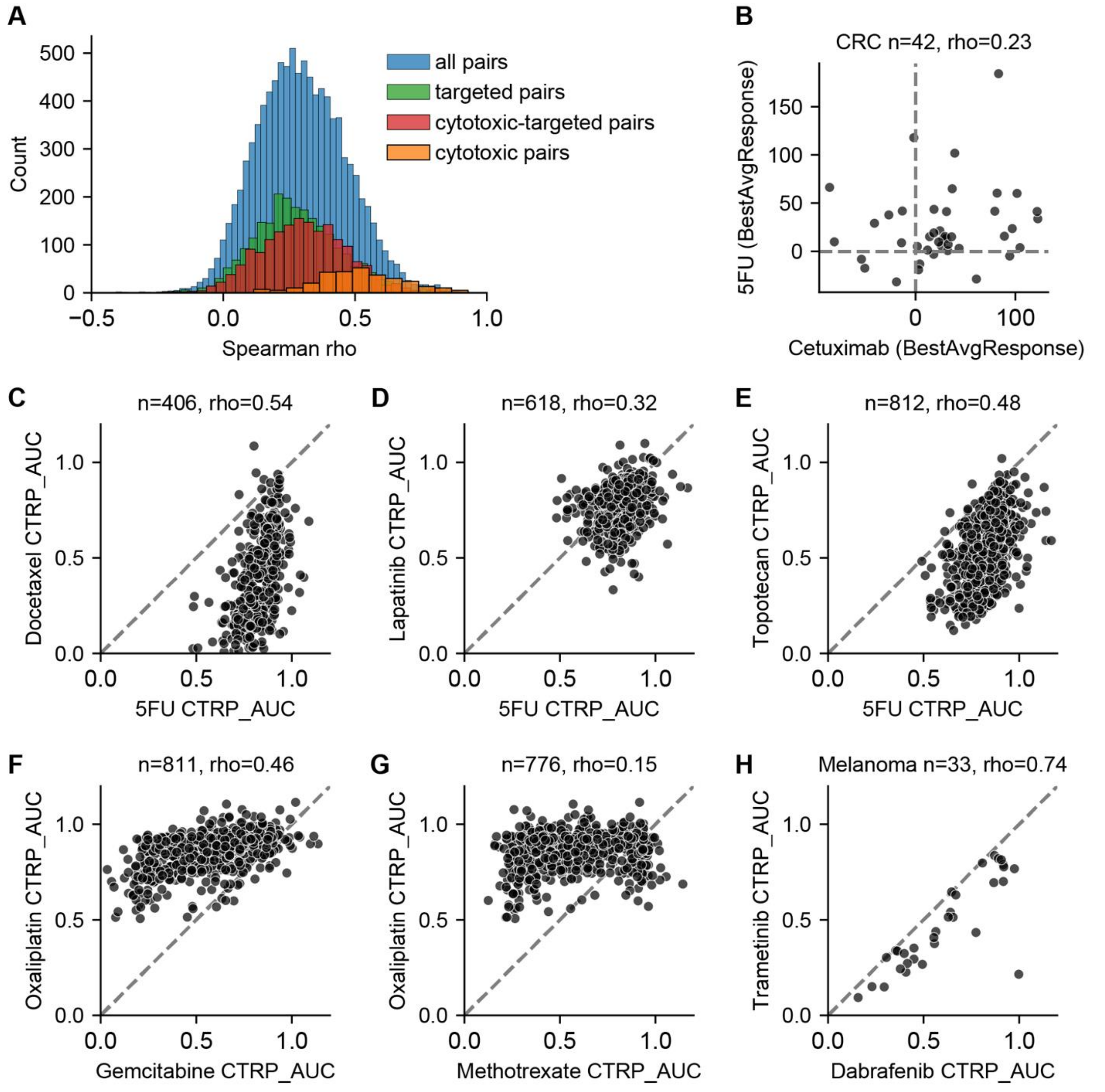


**Extended Data Figure 9**. **Correlations between drug responses from preclinical models were used to compute expected PFS distributions.** (A) Distributions of pairwise Spearman correlations between anti-cancer agents from CTRPv2. (Mean of all drug pairs, 0.30; targeted therapies, 0.28; cytotoxic chemotherapy – targeted therapy pairs, 0.31; cytotoxic chemotherapies, 0.52) (B) Correlation between colorectal cancer PDXs' best average response from 5-fluorouracil (5FU) and cetuximab. Spearman correlations were measured in pan-cancer cell lines for (C) docetaxel and 5FU (substitution for capecitabine), (D) lapatinib and 5FU (substitution for capecitabine), (E) topotecan (substitution for irinotecan) and 5FU, (F) oxaliplatin (substitution for cisplatin) and gemcitabine, and (G) oxaliplatin (substitution for cisplatin) and methotrexate (substitution for pemetrexed). (H) Correlation between trametinib and dabrafenib in melanoma cell lines.

### Extended Data Tables

**Extended Data Table 1**. FDA-approved combination therapies clinical trial data sources and analysis results[4,10–36].
**Extended Data Table 2**. Excluded combination therapies.
**Extended Data Table 3**. Positive and negative phase III trials of combination therapies between 2014 and 2018[22,37–61,61–69].
