## Supplementary Notes for "Additivity predicts the efficacy of most approved combination therapies for advanced cancer"

#### Mathematical derivation of the connection between Bliss independence and addition of PFS times accompanying Supplementary Figure 1.

Our model of clinical drug additivity is formally defined as the addition of PFS times. This null hypothesis does not depend on assumptions about underlying tumor growth kinetics and cytotoxic mechanisms. However, mathematically, the addition of PFS times conveniently corresponds to the Bliss independence model under simplifying assumptions. Bliss independence is a common pre-clinical null-hypothesis used to classify drug synergy or antagonism.

Disease progression is defined as "at least 20% increase in the sum of diameters of target lesions" according to RECIST1.1[1]. Assuming a spherical shape, this corresponds to a ~72% increase in tumor volume or the number of cells. For simplicity, we will define time to disease progression *t*, or progression-free survival, as time to tumor doubling. We will also assume that the tumor grows exponentially. For an untreated tumor with a starting population of *N* and a growth rate of *g*, the time to progression, $PFS_{\text{untreated}}$, can be calculated as:

$$2N = Ne^{gt}$$

$$PFS_{\text{untreated}} = t = \frac{\log 2}{g}$$

Similarly, for a tumor treated with a drug X that reduces the cancer cell population to *xN* (where *x* is the tumor cell fraction that survives therapy), the time to progression after treatment, $PFS_x$, can be calculated as:

$$2N = xNe^{gt}$$

$$PFS_X = t = \frac{\log 2 - \log x}{g}$$

Bliss independence states that for an additive combination A+B, each drug A and B provides a statistically independent probability of cell killing[2]. Thus, the surviving population of cancer cells treated with A+B under Bliss independence will be *abN*, where *a* and *b* are the tumor cell fractions that survive drugs A and B, respectively. Therefore, the time to progression after combination therapy, $PFS_{\text{AB}}$, will be:

$$PFS_{\text{A+B}} = \frac{\log 2 - \log ab}{g}$$

$$= \frac{\log 2 - \log a - \log b}{g}$$

$$= \frac{\log 2 - \log a}{g} + \frac{\log 2 - \log b}{g} - \frac{\log 2}{g}$$

$$= PFS_{\text{A}} + PFS_{\text{B}} - PFS_{\text{untreated}}$$

### Frequently Asked Questions and Answers

This note answers several questions about the model of drug additivity and its implications.

#### How is censoring of patient data addressed?

The Kaplan-Meier method estimates the survival function from censored survival time data. The model of additivity takes as its input published Kaplan-Meier curves of progression-free survival for control arms and novel single therapies. Thus, the effect of censoring has already been accounted for in the publications of these clinical trial results, by their authors' use of the Kaplan-Meier estimator to calculate survival functions from patient data. If one wished to apply the additivity model beginning with individual patient data, rather than a Kaplan-Meier curve, then one should also first calculate the Kaplan-Meier estimated survival function for PFS, then use this as model input. Censoring in data has the effect of decreasing the effective sample size and thereby increasing confidence intervals; with this in mind we elected to not analyze some combinations because sample sizes were small. The clinical trials we analyzed had an average number of participants of 245 per arm, with a range from 41 to 650. For the application of the Cox proportional hazard model, we reconstructed the individual patient events (i.e., disease progression or death, and censoring) from the Kaplan-Meier estimates and the at-risk tables, using published methods[3,4].

#### Why was PFS used as a sole endpoint?

In this work, we analyzed progression-free survival (PFS) as a clinical measurement of anti-cancer drug efficacy. PFS and overall survival (OS) are both used as primary endpoints in phase III clinical trials. OS is affected by subsequent therapies, cross-over, and post-progression survival, whereas PFS is a more direct measure of the assigned treatment's anti-cancer efficacy. While OS is of course a valid trial endpoint (cross-over and subsequent therapies, etc, matter to patient survival and well-being), it is not suited to analyze additive or non-additive effects of drug combinations.

Objective response rate (ORR) and best tumor volume change are used for earlier phase clinical trials but are not primary endpoints for pivotal phase III clinical trials. The HSA model (termed as the independent drug action model in previous work) can be applied to tumor volume change and ORR, as reported by ourselves and others[8–10]. However, because an objective response is a categorical variable (a given patient either has a response or does not), there is not yet a clear basis for defining 'additivity' in ORR or volume change. As described below, the sum of response probabilities is not a biologically or mathematically plausible model and is profoundly different from the model presented here of adding PFS durations.

#### How does the model address toxicity, dosing schedule, and dose reductions?

This work does not attempt to predict toxicity. In general, the FDA-approved combinations we have analyzed are tolerable combinations. We have only analyzed combinations where the constituent drugs were administered at the same dose as monotherapy datasets or at a minimum of 75% of the monotherapy dose (**Supplementary Figure 2**). Specifically, 29 of 37 combinations used 100% of monotherapy doses, one used 112% of monotherapy doses, and 7 of 37 combinations used between 75% and 90% of monotherapy doses. We cannot rule out the possibility of enough synergy to balance a 10% to 25% dose adjustment. Administration schedules were similar between combination therapy and matched monotherapy data (**Supplementary Table 1**). Trials may have had differences between arms in dose modifications or interruptions after commencing therapy, but in general, do not report this information in sufficient detail to assess its potential impact.

Our clinical additivity model was based on 'effect addition' (which has a pre-clinical analogy in Bliss independence). We did not attempt to expand our model to 'dose addition' (as in Loewe additivity) because phase III clinical trials do not measure dose-response relationships in patients. For this reason, we have declined to analyze combinations with significant dose reductions. A typical case that we have declined to analyze is when one drug in the combination is administered at a lower dose than was used in its monotherapy studies. If a combination is administered with one agent at lower dose, and its efficacy is consistent with drug additivity assuming full dose, then two competing hypotheses could explain the observation:

(1) The single-agent's efficacy is not diminished with lowered dose, and the effect of the combination is additive.

(2) The combination is synergistic (by Loewe's model, i.e., potency is enhanced).

These two explanations are indistinguishable without data on the dose-response relationship in humans. Clinically administered doses are often set at the maximum tolerated dose identified in phase I trials, and for many cancer therapies, clinical studies have observed that lowering a drug's dose as much as two-fold does not diminish its clinical efficacy[5–7]. For this reason hypothesis (1) is a possibility unless one has data on single-agent efficacy at the relevant dose.

Similarly, measurements of drug effect at ill-matched doses prevents the ability to assess drug interactions even in cell culture experiments (**Fig. S1**). Effect addition (Bliss independence) can be only tested when the combination is at the full dose (**Fig. S1A**). Dose addition (Loewe additivity and combination index) needs to be tested along the equipotent line, depicted as the purple dotted line in Fig. S1B. Even one measurement of the combination activity along this line can determine if there is 'excess over additivity'. However, this cannot be determined when the combination is only measured outside this line. The excluded combination therapies all fall into this inadequate dose range, and thus, for the same reasons, it is not possible to assess whether they are interacting or non-interacting in either the 'Bliss independence' or 'Loewe additivity' sense.

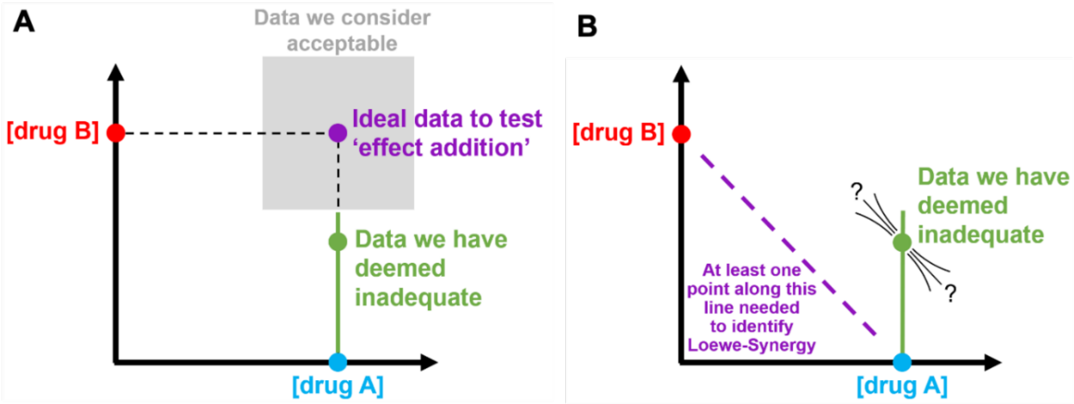


**Figure S1. Combinations with reduced doses are mostly inadequate to assess drug interactions.** (A) The ideal set of measurements to assess effect addition is when two drugs are combined at full dose. The combination therapies analyzed here were administered at least 75% of the monotherapy doses. (B) To test Loewe's dose-additivity model, at least one measurement of combination activity should be along the equipotent line. In the combination therapies we have declined to analyze, the available data cannot determine the shape of the isobole and therefore cannot test whether a combination is Loewe-synergistic.

**Should these human clinical trial results be verified in a mouse?**

The results of randomized, controlled phase III clinical trials in humans are in general a higher standard of evidence than mouse experiments. However, there are also scientific reasons why key features of combination therapy in diverse human populations are not reproduced using replicates of homogenous mouse models (even if a mouse was a perfect model of therapeutic effect in a human). A key feature of this model of drug additivity lies in recognizing that cancer therapies have variable efficacy among human patients, and therefore combinations of therapies consist of adding a variable effect to another variable effect (**Figure 1A**). The variation among patients in the best single drug response (HSA model) is quantitatively the largest contributor to the benefit of most drug combinations. Because the variable efficacy in humans is not apparent in replicates of any one mouse model, this major contributor to the benefit of drug combinations is fundamentally not observable in a study on replicates of a single mouse model. A more human-relevant view may be provided by using heterogeneous panels of patient-derived tumor xenografts (PDXs) as models of heterogeneous populations of human cancers. We previously analyzed ~4,500 PDX experiments and observed that patient-to-patient variability in drug response quantitatively explains a majority of the benefit of many combination therapies as observed in PDXs[8]. Thus, the phenomenon in general has been verified in thousands of PDX experiments.

A final consideration is that mouse experiments assessing combinations of cancer therapies commonly test whether combination therapy elicits longer survival or greater tumor shrinkage compared with monotherapy. This is a justified endpoint for anticipating clinical potential, but it is not a test of drug interaction, because observed efficacy was not compared to a calculated expectation of drug additivity (non-interaction). Therefore, mouse experiments on combination therapies generally have not tested whether combination therapy is synergistic, additive, or less than additive. Note, our analysis does not challenge the utility of mouse experiments, because a general conclusion of this study is that a 'more than additive' effect is not necessary for clinical benefit. These considerations do however highlight that conclusions about drug synergy from mouse experiments are routinely unjustified: it is erroneous to conclude that all combination therapies which are superior to monotherapy have a synergistic drug-drug interaction.

**Why isn't the sum of probabilities used as a model of drug additivity?**

The arithmetic addition of probability densities is mathematically and biologically nonsensical; it cannot describe even the simple scenario of rolling two dice (**Figure 1A**). Consider two therapies that each elicit PFS times between 1 and 10 months, with a 10% chance of each (akin to a 10-sided die). If the sum of probabilities were used as a model, it would predict a 20% chance of each outcome (being PFS = 1 month, 2 months, etc.); all possible outcomes have a total probability of 200%. It should be apparent that the total probability will be 200% for any distribution of outcomes, which is mathematically nonsensical. Summing survival functions (1 minus the cumulative density function) is no better, as survival at time 0 would be 200%. In a regime where single agent PFS probabilities are below 50%, summing probabilities is still not biologically plausible. For example, if two single agents each elicit 30% PFS at 12 months, then a model of 30% + 30% = 60% PFS describes a scenario where no patients at all would be responsive to either therapy. Clinical and experimental data from sequentially treated patients, or patient-derived tumor xenografts tested against multiple therapies, all show that some tumors are sensitive to more than one therapy[8,11], thus such a model is inappropriate. (This article's model of adding PFS times accounts for tumors being responsive to multiple therapies). Conversely, one could consider multiplying failure rates (e.g. 50% and 50% PFS are predicted to produce 75% PFS); this corresponds to the model of Highest Single Agent and does not describe an additive effect of multiple drugs in individual patients[8]. Thus, for many mathematical and biological deficiencies, summing probabilities is not a viable model of drug additivity.

**Why isn't the product of hazard ratios used as a model of drug additivity?**

This question asks whether drug additivity can be defined as multiplying Hazard Ratios (HR), such that HR(A+B) = HR(A) × HR(B), which corresponds to the sum of each drug's log hazard ratio (log(HR(A+B)) = log(HR(A)) + log(HR(B))). Hazard ratio is a relative quantity that compares the hazard of a treatment to a comparator treatment (or lack of treatment). One initial problem with this approach is that different trials may use different comparators (control arms), making their hazard ratios not directly comparable. However, a more severe problem is that for any drugs with defined efficacy, this definition makes different predictions based on the choice of the comparator. This can be illustrated by an exponential survival function. Suppose a common comparator (therapy X) has a hazard of 0.5 (survival function = exp(-0.5 t)), and treatments A and B confer hazards of 0.2 and 0.25, respectively (survival functions = exp(-0.2 t) and exp(-0.25 t), respectively). Then treatments A and B have Hazard Ratios of 0.4 and 0.5 versus treatment X, and the predicted effect of combination A+B is a Hazard Ratio of 0.4 × 0.5 = 0.2 versus treatment X (survival function = exp(-0.1 t)). However, if the control arm had been a placebo with a hazard of 1.0 (i.e, inferior to treatment X), then treatments A and B, though there is no change in their survival functions, would be assigned different hazard ratios, being 0.2 and 0.25 versus placebo. In this case, the predicted effect of combination A+B is a hazard ratio of 0.2 × 0.25 = 0.05 versus placebo (survival function = exp(-0.05 t)). Thus, the product of hazard ratios makes different predictions based on the choice of comparator, even with no change in the activity of the drugs comprising a combination. Even were a consistent placebo used as the comparator for such calculations, it would often make absurdly optimistic predictions, such as a Hazard Ratio of 0.05 in the example above. This results in PFS times that are much more than arithmetically additive (**Figure S1**). Thus, the product of hazard ratios is not a viable model of drug additivity because its predictions would be mathematically inconsistent and biologically unrealistic.

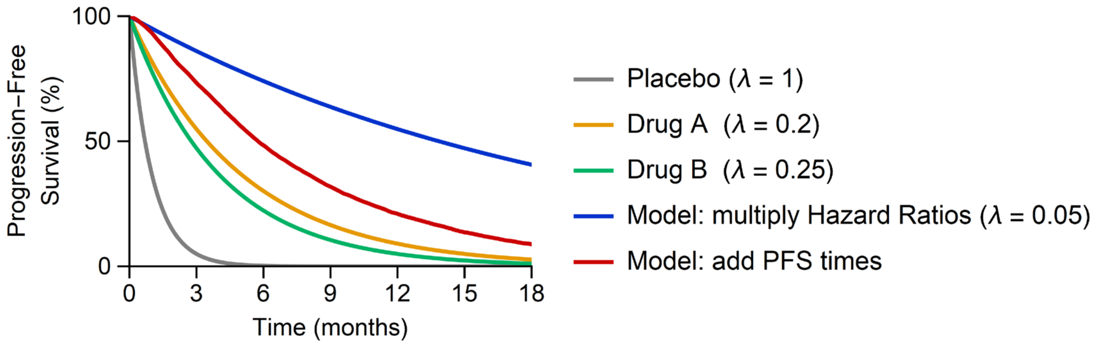


**Figure S2. Multiplying Hazard Ratios results in unrealistic predictions with survival times that are far longer than arithmetic addition.** Exponential survival functions were simulated with rate parameters of $\lambda = 1$, 0.2, and 0.25, for placebo, drug A, and drug B, respectively. The effects of an 'additive' combination were simulated by the addition of PFS times (the model proposed in this study, using correlation $\rho = 0.3$ and scan interval 1.5 months), or by multiplying the hazard ratios of single drugs relative to placebo, which results in a rate of $\lambda = 0.05$. Median PFS time from multiplying hazard ratios is four times as large as the most active single drug (14 months vs. 3.5 months), making it much more than the addition of single drug PFS times.

**How do HSA and additivity models work when adding two wholly ineffective therapies?**

Combining wholly ineffective therapies should also result in wholly ineffective combination therapy when assuming additivity. Wholly ineffective therapy is equivalent to treating cancers with a placebo or the best supportive care. As illustrated in **Supplementary Fig. 3**, advanced cancers progress within the first few months when receiving no anti-cancer therapy (most tumors exhibit progression before or on their first scheduled radiological scan). We have previously shown that the HSA model does not predict improvement when combining two placebos; across many clinical datasets of placebo-only treatment, the expected benefit of placebo plus placebo is 0% to 1.5% survival at 12 months[12]. The additivity model makes a similar prediction, with near zero benefits when combining two placebo drugs (**Fig. S2**).

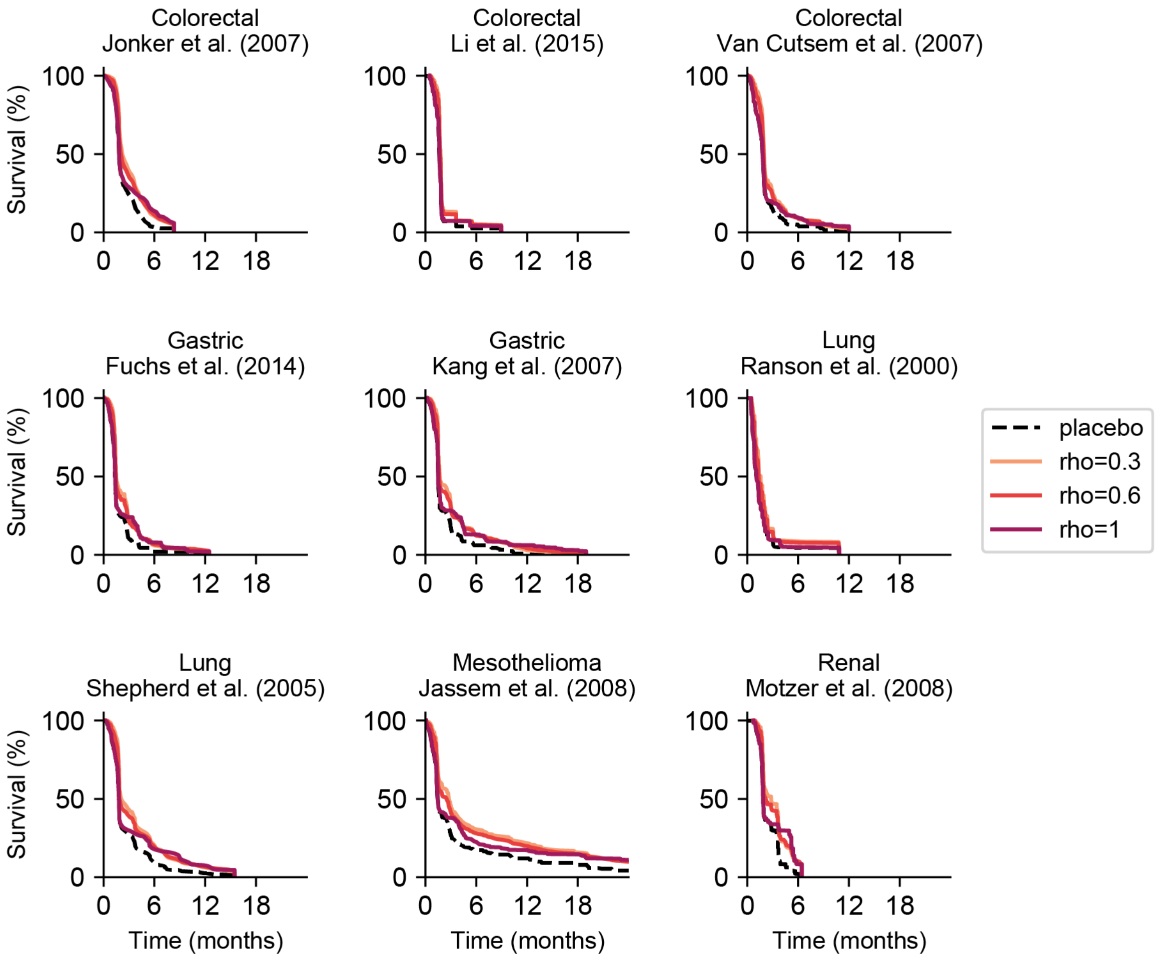


**Figure S3. The additivity model predicts near zero benefit when combining placebo or best supportive care (i.e., wholly ineffective anti-cancer) drugs.** Since wholly ineffective drugs will have the same effect (i.e., no therapeutic benefit) on patients, the correlation in response should be $\rho = 1$. We have also included hypothetical cases where the correlation is lower to demonstrate that the level of correlation has a small impact on the predictions in this scenario.

**Would drugs with low correlation in drug responses (low cross-resistance) have better efficacies under the additivity model?**

Whereas the HSA model is influenced by the correlation between monotherapy responses across a patient population, correlation has very little influence on the combination's efficacy under the additivity model (**Fig. S4**). The benefit of the HSA model entirely relies on the inter-patient heterogeneity between drug responses. For the additivity model, the same quantities are being added irrespective of the level of correlation, and thus, the average efficacy of the combination remains the same. This is the nature of the sum of two random variables: $E(X) + E(Y) = E(X+Y)$. The variance of the distribution, on the other hand, is affected by correlation as $Var(X+Y) = Var(X) + Var(Y) + Cov(X, Y)$. A low correlation will yield lower variance, and a high correlation will yield higher variance. This can have a minor impact on the effect size as measured by the hazard ratio from the Cox proportional hazard model (**Fig. S4**), where a higher correlation will result in a small increase in the hazard ratio (i.e., less benefit).

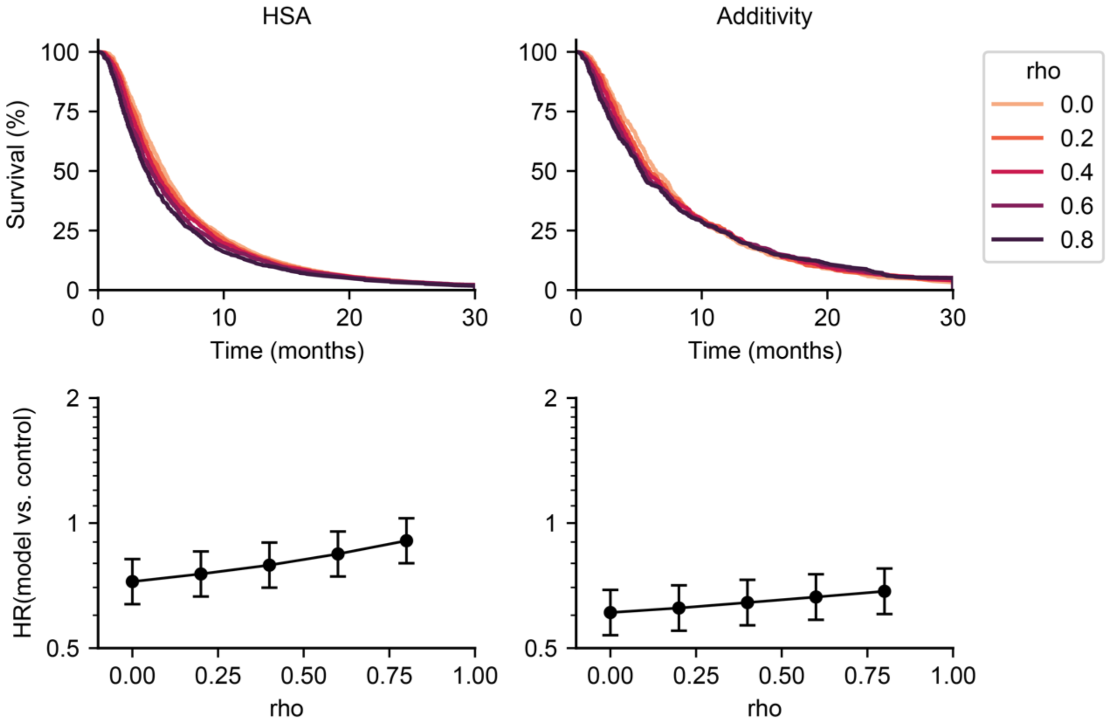


**Figure S4. Additivity is less affected by the correlation of monotherapy responses compared to HSA.** Combination effects under the HSA model (top left) and the additivity model (top right) vary modestly by the level of correlation (rho) between the monotherapy drug responses. Here, examples of monotherapy PFS distributions were constructed from lognormal survival functions, with an experimental drug ($\mu$=1, $\sigma$=1, natural log base) and control drug ($\mu$=1.2, $\sigma$=1, natural log base). Either the HSA model (bottom left) or the additivity model (bottom right) was compared to the control drug by the Cox proportional hazard model. Under the HSA model, combination efficacy was modestly improved by lower correlation, but under the additivity model, correlation has negligible influence. Error bars indicate 95% confidence intervals.
